# Global Detection of Respiratory Illness Outbreaks in Travelers: A Statistical Approach using GeoSentinel Data, 2015–2024

**DOI:** 10.64898/2026.05.06.26352534

**Authors:** Stan Heidema, Ivo V. Stoepker, Daniel T. Leung, Watcharapong Piyaphanee, Lin H. Chen, Marta Díaz-Menéndez, Kevin O’Laughlin, Michael Libman, Davidson H. Hamer, Edwin R. van den Heuvel, Ralph Huits

## Abstract

**Background:** Novel respiratory pathogens have pandemic potential, making epidemiologic surveillance of acute lower respiratory tract infections (acute LRTI) a global public health priority. Monitoring acute LRTI among international travelers provides an underutilized opportunity to complement existing surveillance systems, though reliable denominator data on travel volume are often unavailable.

**Aim:** We aimed to develop and validate a framework for detecting acute LRTI outbreaks among international travelers in the absence of reliable denominator data.

**Methods:** Using syndromic and etiologic GeoSentinel data from 2015 to 2019, we modeled baseline LRTI epidemiology in travelers by comparing generalized linear mixed models (GLMMs) and selecting the preferred model using out-of-sample metrics. A robust Shewhart control-chart framework, accounting for increases in travel volume under non-epidemic conditions, was applied to detect deviations from expected trends and retrospectively to 2020 data from 64 countries to identify early COVID-19 signals.

**Results:** The preferred hybrid autoregressive GLMM, incorporating country-specific fixed effects, random seasonal effects, and a latent temporal autocorrelation structure, demonstrated adequate goodness-of-fit across pre-pandemic and post-pandemic (2023 to 2024) periods. The framework detected an early syndromic signal in China under the conservative assumption of up to a threefold increase in travel volume, consistent with COVID-19 emergence; a signal was also detected in Italy, driven primarily by influenza rather than novel syndromic cases.

**Conclusion:** Combining traveler surveillance with this statistical framework may support early detection of acute LRTI outbreaks despite absent denominator data, positioning GeoSentinel as a valuable complementary network for global health security.

## 1 Introduction

The pandemic potential of respiratory illnesses, as evidenced by four influenza-induced pandemics in the 20th century and the coronavirus disease 2019 (COVID-19) pandemic in the 21st century demonstrate the need for continuous active and passive surveillance of respiratory pathogens [1]. Many surveillance systems have been developed and currently contribute to pandemic preparedness, such as the Global Influenza Surveillance and Response System (GISRS), established in 1952 which plays a pivotal role in establishing the composition of the yearly influenza vaccine [2].

Sentinel surveillance of international travelers is a cost-effective tool to monitor circulation of pathogens because travelers are frequently exposed to respiratory pathogens that they may then import into their home countries [1]. In addition, travelers may have increased exposure and susceptibility compared to local populations due to increase of time spent in crowded, closed space environments or being immunologically naïve due to seasonality of infections (e.g., influenza, COVID-19). Therefore, traveler surveillance offers a valuable complement to domestic public health activities [3]. Recognizing this, the U.S. Centers of Disease Control (CDC) conducts large-scale traveler surveillance through its Traveler-based Genomic Surveillance (TGS) program [4].

The GeoSentinel network, founded in 1995 by CDC and the International Society of Travel Medicine, is a global surveillance system that collects clinical data on infectious diseases and other adverse health events affecting international travelers and migrants. The network includes approximately 70 member sites across 30 countries in six continents [5], where clinicians specializing in travel and tropical medicine collect data while diagnosing and treating patients. Historically, GeoSentinel has contributed to the evidence base for illnesses that afflict travelers and the detection of outbreaks caused by diverse pathogens [5]. However, its potential for sentinel surveillance of acute respiratory illnesses, whether to detect outbreaks of known pathogens or the emergence of novel respiratory diseases, has been less emphasized.

While established surveillance systems such as the GISRS and, more recently, the TGS program have greatly strengthened global detection of respiratory pathogens through country-level laboratory capacity and structured sampling at ports of entry [4, 6], traveler-based clinical surveillance networks such as GeoSentinel provide an important complementary approach. By collecting detailed syndromic and diagnostic data from returning travelers exposed across diverse settings [5, 7], these systems can detect unusual patterns outside traditional hospital- or country-based frameworks, thereby filling critical gaps in global respiratory pathogen monitoring and contributing to earlier identification of emerging threats.

The primary limitation (see, for example [7, 8]) for the use of GeoSentinel data is the difficulty of obtaining reliable denominator data, i.e., the total number of international travelers who potentially could seek care at a GeoSentinel site when ill (hereafter termed “travel volume” for simplicity). Firstly, the absence of such data complicates the accurate modeling of nonzero baseline epidemiology, a task already recognized as inherently challenging in sentinel surveillance [9, 10]. Secondly, without travel volume data, a surge in cases can be attributed both to an increase in rate of respiratory illness (i.e., an outbreak), as well as to an increase in travel volume.

In this work, we propose an approach that leverages GeoSentinel’s unique dataset to complement existing surveillance efforts by circumventing the explicit need for denominator data, as long as justifiable assumptions can be made about its evolution over time. We illustrate this method by retrospectively identifying potential early COVID-19 signals, as indicated by increased syndromic case counts among international travelers in early 2020.

## 2 Methods

### 2.1 Study design and data source

We analyzed data from the GeoSentinel network database. Clinicians at GeoSentinel’s member sites collect surveillance data from patients using standardized diagnostic codes. Patients are eligible for entry into the GeoSentinel database if they have crossed an international border and are seen at a GeoSentinel site with a possible travel-related illness. The decision to enter a case into the database, including determination of whether the illness is travel-related, is made by the site. A standardized data collection form is used to collect de-identified demographic, clinical, and travel-related information, and data are entered electronically into a secure web-based centralized database. Standardized diagnostic codes corresponding to syndromic and/or etiologic diagnoses, with established and clearly defined case definitions, are entered by the site for each case. The database contains automated procedures to ensure completeness and integrity of data, and GeoSentinel personnel manually evaluate records that are flagged for irregularities by either the database or the site.

A consensus-based approach among clinician co-authors was used to classify GeoSentinel diagnostic codes for acute lower respiratory tract infections (LRTIs). Conditions unanimously included were atypical non-lobar unspecified pneumonia, COVID-19, influenza A, influenza B, influenza-like illness, influenza (avian), Legionnaires’ disease, MERS-CoV, pneumococcal pneumonia, pneumonia (bacterial or viral, other specific etiology), pneumonia lobar unspecified, and severe acute respiratory syndrome (SARS). Conditions excluded as not meeting the agreed criteria were cough without etiology, laryngitis, pharyngitis (streptococcal or unspecified), and upper respiratory infection. We conducted a sensitivity analysis (see Supplementary Material) for codes where expert opinion differed regarding their classification as acute LRTIs: acute bronchitis, acute respiratory distress syndrome (ARDS), and viral respiratory infection of other specific etiology.

By aggregating relevant acute LRTI conditions using a consensus-based approach, we aimed to strike a balance between a fragmented approach—where each condition is monitored separately, requiring conservative thresholds to maintain acceptable control over false positives—and excessive aggregation, which risks masking smaller, condition-specific outbreaks. While the included diagnoses were etiologically distinct, their clinical presentations overlapped within syndromic categories. Considering these collectively as acute LRTI within the model captured both confirmed and syndromic cases, providing robustness to variability in reporting practices and enabling detection of outbreaks from both known and novel respiratory pathogens.

Analyzed data included country of exposure and clinic visit date. We obtained acute respiratory illnesses from the GeoSentinel database from January 1, 2015, through December 31, 2024. We used records from 2015–2019 for baseline model development and validation, 2020 data for retrospective COVID-19 outbreak detection, and 2023–2024 data to assess post-pandemic baseline model performance. Data from 2021–2022 were excluded because international travel was heavily restricted during this period, resulting in case volumes too low to be representative.

Individual conditions contributing fewer than 10 yearly cases worldwide during the baseline period were excluded, as the proposed methodology was not reliable for such rare conditions: their low frequency rendered them highly susceptible to masking by higher-frequency diseases when aggregated. As an important consequence of this exclusion criterion, diagnostic codes that were added only after the baseline period ended were also excluded; most notably COVID-19. This allowed us to illustrate how—when deployed with data that was available in real time only—the proposed framework could detect outbreaks caused by (at the time of surveillance) unknown pathogens, through manifestation of an increase in related LRTI case counts. Countries of exposure with fewer than two acute LRTI cases per year on average during the baseline period were also excluded, as the models required sufficient case volume for reliable baseline parameter estimation.

### 2.2 Modeling baseline epidemiology: generalized linear mixed models

To detect abnormal events, we established a baseline expectation by analyzing weekly case counts for each country of exposure. This process modeled the typical epidemiological pattern in the absence of an outbreak, which is an important prerequisite for outbreak detection systems [9, 10]. Because case numbers were expected to vary substantially between countries and to exhibit country-specific yearly seasonality—given the seasonal nature of both respiratory illness transmission [1] and international travel volumes [11]—we employed a generalized linear mixed model (GLMM) with a negative binomial distribution. Specifically, we modeled the aggregated weekly number of acute LRTIs *Y*_*it*_ for country of exposure *i* at week index *t* as *Y*_*it*_ | *µ*_*it*_, *θ* ~ NegBin(*µ*_*it*_, *θ*), where *θ* quantifies the overdispersion of weekly counts. Our primary approach, the hybrid autoregressive model, accounts for seasonality by treating country-specific baselines as fixed effects and seasonal deviations as random effects following a sinusoidal pattern. To capture the temporal dependency inherent in infectious disease data, we incorporated a latent temporal autocorrelation term *ϵ*_*it*_ into the mean model:

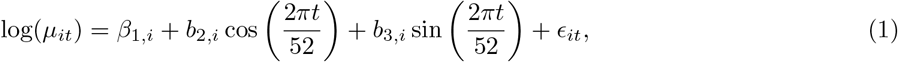

where the seasonal coefficients *b*_2,*i*_ and *b*_3,*i*_ follow independent normal distributions. The term *ϵ*_*it*_ is normally distributed and reflects a periodic temporal autocorrelation structure to account for the annual cycle:

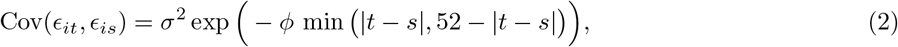

where *t* and *s* are week indices, and observations from different countries of exposure remain uncorrelated (Cov(*ϵ*_*it*_, *ϵ*_*js*_) = 0 if *i* ≠ *j*). We compared this hybrid autoregressive model with six alternative approaches to validate its effectiveness in modeling baseline epidemiology: full fixed-effects, full random-effects, hybrid-effects (without autocorrelation), direct clustering, spherical clustering, and an autoregressive model (without harmonic terms). We evaluated model generalizability through out-of-sample validation using log-likelihood, root-mean-square error (RMSE), log score (LogS), and continuous ranked probability score (CRPS) [12]. We examined the hybrid autoregressive model’s absolute performance through in-sample diagnostics for the nine countries with the highest case counts. This included visual inspection of fitted values [12, 13], randomized quantile residual analysis [14–16], and inspection of residual temporal and spatial autocorrelations. All statistical analyses were performed using R version 4.2.3 [17]. Further details on model specification and implementation are provided in the Supplementary Material.

### 2.3 Outbreak detection: robust Shewhart charts

After establishing baseline behavior by obtaining estimates 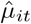 and 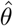, weekly case counts were monitored using Shewhart control charts [18], comparing observed values with expectations derived from the hybrid autoregressive model. Because *µ*_*it*_ approximates the product of *individual risk* and *travel volume*, representing the expected number of travelers seeking care at a GeoSentinel site when ill, a proportional change *c* in travel volume scales the mean to *cµ*_*it*_. To make the approach robust against signals driven by increases in travel volume rather than individual risk, we assumed a maximum allowable multiplicative increase in travel volume under non-epidemic conditions, denoted by *c* (*c* = 1, 2, or 3). We therefore defined the null hypothesis as 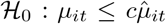, so that higher values of *c* corresponded with a more conservative design, requiring a larger observed case count to rule out increased travel volume as an alternative explanation for the signal. A signal was triggered when the observed number of cases exceeded the (1 − *α*) quantile of the fitted negative binomial distribution. We set *α* so that, globally, we expected approximately one false signal every 156 weeks (about three years), a frequency deemed manageable by travel epidemiologists. To achieve this, we applied a Bonferroni correction and, for 64 countries, set *α* = 1.0016 × 10^−4^, which yielded an in-control average run length (ARL_0_) of approximately 10,000 weeks (about 192 years) at the individual country level, reflecting a highly conservative false-alarm rate. When a signal occurred, we reviewed the corresponding code categories to determine whether it reflected known seasonal infections (e.g., influenza) or indicated potentially novel respiratory events without an identified pathogen. We retrospectively applied the outbreak-detection framework to early 2020 data to assess its capacity to detect signals related to the emergence of COVID-19.

## 3 Results

### 3.1 Data description

Table 1 summarizes the number of reported cases across the study periods. We retrieved 5,578 records of acute lower respiratory illness from the GeoSentinel database, of which 661 were excluded based on the described criteria. Most excluded cases (*n* = 639) were COVID-19 diagnoses from 2020, which were removed because this condition did not contribute cases during the 2015–2019 baseline period and therefore failed to meet the predefined selection criterion. The final dataset included 4,917 cases and covered 64 countries in 2015–2019 and 2020, and 63 countries in 2023–2024, distributed globally across all continents except Antarctica. This led to *n* = 3, 478 cases from 2015–2019, *n* = 284 from 2020, and *n* = 1, 115 from 2023–2024. Across all included cases, *n* = 2, 528 (51.4%) were etiologic diagnoses and *n* = 2, 389 (48.6%) were syndromic. Influenza A was the most frequent etiologic condition (*n* = 1, 219), and influenza-like illness was the leading syndromic category (*n* = 1, 447).

**Table 1.** Reported acute lower respiratory illness cases by condition and inclusion status, across 64 countries (2015–2019), 64 countries (2020), and 63 countries (2023–2024). Conditions for which consensus was not reached, conditions contributing fewer than 10 yearly cases globally during the baseline period, and countries with fewer than 2 average annual acute LRTI cases were excluded.

| **Included Conditions** |  |  |  |  |  |
| --- | --- | --- | --- | --- | --- |
| **2015–2019 (64 countries)** | *n* | **2020 (64 countries)** | *n* | **2023–2024 (63 countries)** | *n* |
| *Etiologic* |  |  |  |  |  |
| Legionnaires' Disease | 45 | Legionnaires' Disease | 2 | COVID-19 | 419 |
| Influenza A | 830 | Influenza A | 101 | Influenza A | 288 |
| Influenza B | 351 | Influenza B | 37 | Influenza B | 93 |
| Pneumonia (bacterial or viral, other specific etiology) | 272 | Pneumonia (bacterial or viral, other specific etiology) | 21 | Pneumonia (bacterial or viral, other specific etiology) | 69 |
| *Syndromic* |  |  |  |  |  |
| Influenza-like illness | 1235 | Influenza-like illness | 78 | Influenza-like illness | 134 |
| Pneumonia, atypical/non-lobar, unspecified | 255 | Pneumonia, atypical/non-lobar, unspecified | 23 | Pneumonia, atypical/non-lobar, unspecified | 42 |
| Pneumonia, Lobar, Unspecified | 490 | Pneumonia, Lobar, Unspecified | 22 | Pneumonia, Lobar, Unspecified | 110 |
| **Total (Included)** | **3478** | **Total (Included)** | **284** | **Total (Included)** | **1155** |
| **Excluded Conditions** |  |  |  |  |  |
| *Etiologic* |  |  |  |  |  |
| COVID-19 | 0 | COVID-19 | 639 | Legionnaires' Disease | 3 |
| Influenza, Avian | 0 | Influenza, Avian | 0 | Influenza, Avian | 0 |
| MERS-Co-V | 0 | MERS-Co-V | 0 | MERS-Co-V | 0 |
| SARS | 0 | SARS | 0 | SARS | 0 |
| *Syndromic* |  |  |  |  |  |
| Pneumonia, pneumococcal | 7 | Pneumonia, pneumococcal | 3 | Pneumonia, pneumococcal | 9 |
| **Total (Excluded)** | **7** | **Total (Excluded)** | **642** | **Total (Excluded)** | **12** |
| **Total** | **3485** | **Total** | **926** | **Total** | **1167** |

### 3.2 Comparison of models for baseline epidemiology

We evaluated out-of-sample performance using Log-Likelihood, RMSE, LogS, and CRPS metrics for seven candidate models: full fixed-effects, full random-effects, hybrid, direct clustering, spherical clustering, autoregressive, and the hybrid autoregressive model (Table 2). Overall, the hybrid autoregressive model performed best on average across all performance metrics, indicating that it was the most suitable option for modeling baseline epidemiology.

**Table 2.**
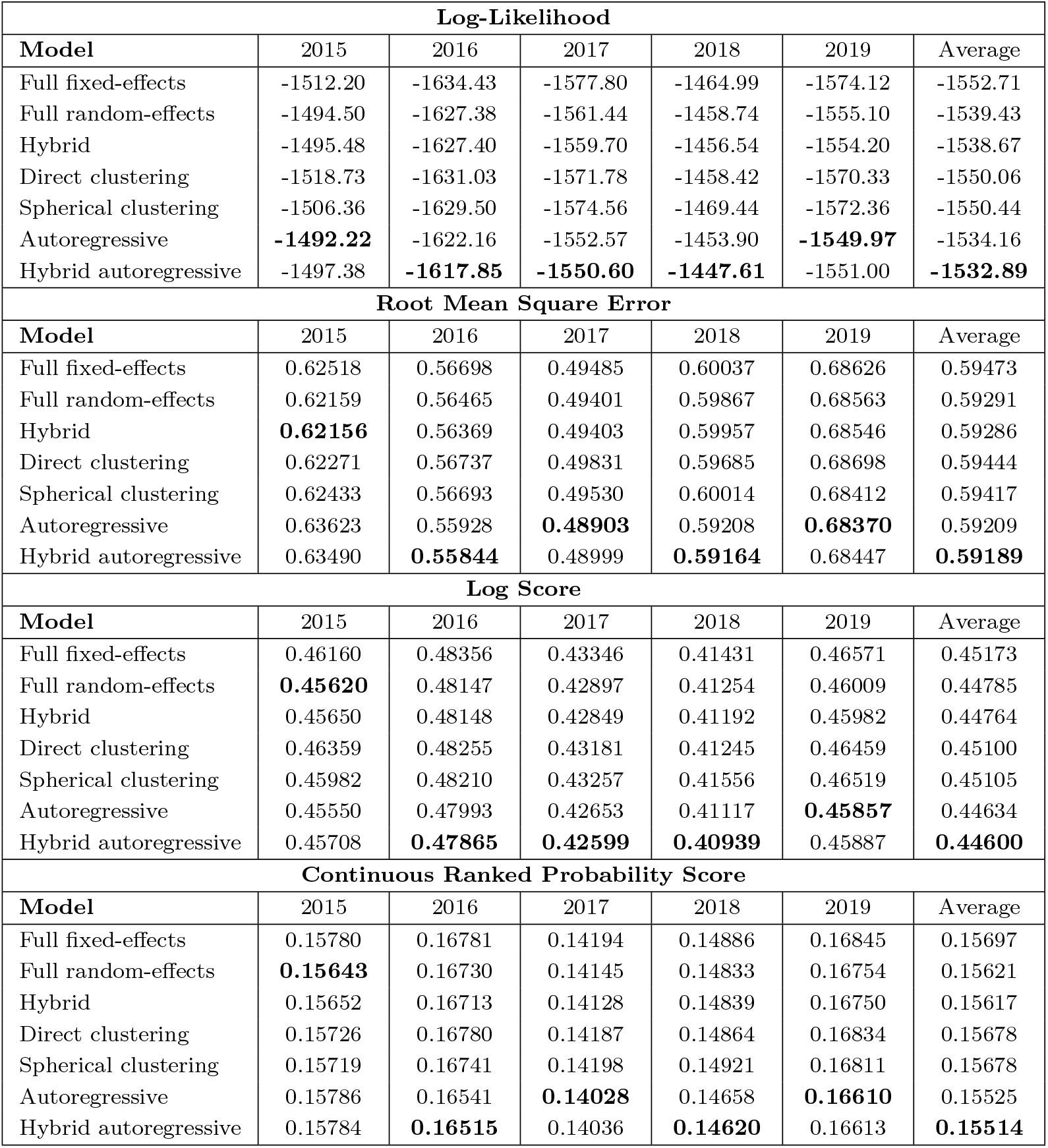
Out-of-sample performance metrics (Log-Likelihood, Root Mean Square Error (RMSE), Log Score (LogS), and Continuous Ranked Probability Score (CRPS)) are presented for seven candidate models (full fixed-effects, full random-effects, hybrid, direct clustering, spherical clustering, autoregressive, hybrid autoregressive). Each column indicates the out-of-sample year, where that year’s data were excluded from training and only used for assessment. The ‘Average’ column represents the mean performance across all out-of-sample years. Higher Log-Likelihood values indicate better performance, while lower RMSE, LogS, and CRPS scores indicate better performance. Bold values highlight the best performance for each column within each metric.

| Log-Likelihood |  |  |  |  |  |  |
| --- | --- | --- | --- | --- | --- | --- |
| Model | 2015 | 2016 | 2017 | 2018 | 2019 | Average |
| Full fixed-effects | -1512.20 | -1634.43 | -1577.80 | -1464.99 | -1574.12 | -1552.71 |
| Full random-effects | -1494.50 | -1627.38 | -1561.44 | -1458.74 | -1555.10 | -1539.43 |
| Hybrid | -1495.48 | -1627.40 | -1559.70 | -1456.54 | -1554.20 | -1538.67 |
| Direct clustering | -1518.73 | -1631.03 | -1571.78 | -1458.42 | -1570.33 | -1550.06 |
| Spherical clustering | -1506.36 | -1629.50 | -1574.56 | -1469.44 | -1572.36 | -1550.44 |
| Autoregressive | **-1492.22** | -1622.16 | -1552.57 | -1453.90 | **-1549.97** | -1534.16 |
| Hybrid autoregressive | -1497.38 | **-1617.85** | **-1550.60** | **-1447.61** | -1551.00 | **-1532.89** |
| Root Mean Square Error |  |  |  |  |  |  |
| Model | 2015 | 2016 | 2017 | 2018 | 2019 | Average |
| Full fixed-effects | 0.62518 | 0.56698 | 0.49485 | 0.60037 | 0.68626 | 0.59473 |
| Full random-effects | 0.62159 | 0.56465 | 0.49401 | 0.59867 | 0.68563 | 0.59291 |
| Hybrid | **0.62156** | 0.56369 | 0.49403 | 0.59957 | 0.68546 | 0.59286 |
| Direct clustering | 0.62271 | 0.56737 | 0.49831 | 0.59685 | 0.68698 | 0.59444 |
| Spherical clustering | 0.62433 | 0.56693 | 0.49530 | 0.60014 | 0.68412 | 0.59417 |
| Autoregressive | 0.63623 | 0.55928 | **0.48903** | 0.59208 | **0.68370** | 0.59209 |
| Hybrid autoregressive | 0.63490 | **0.55844** | 0.48999 | **0.59164** | 0.68447 | **0.59189** |
| Log Score |  |  |  |  |  |  |
| Model | 2015 | 2016 | 2017 | 2018 | 2019 | Average |
| Full fixed-effects | 0.46160 | 0.48356 | 0.43346 | 0.41431 | 0.46571 | 0.45173 |
| Full random-effects | **0.45620** | 0.48147 | 0.42897 | 0.41254 | 0.46009 | 0.44785 |
| Hybrid | 0.45650 | 0.48148 | 0.42849 | 0.41192 | 0.45982 | 0.44764 |
| Direct clustering | 0.46359 | 0.48255 | 0.43181 | 0.41245 | 0.46459 | 0.45100 |
| Spherical clustering | 0.45982 | 0.48210 | 0.43257 | 0.41556 | 0.46519 | 0.45105 |
| Autoregressive | 0.45550 | 0.47993 | 0.42653 | 0.41117 | **0.45857** | 0.44634 |
| Hybrid autoregressive | 0.45708 | **0.47865** | **0.42599** | **0.40939** | 0.45887 | **0.44600** |
| Continuous Ranked Probability Score |  |  |  |  |  |  |
| Model | 2015 | 2016 | 2017 | 2018 | 2019 | Average |
| Full fixed-effects | 0.15780 | 0.16781 | 0.14194 | 0.14886 | 0.16845 | 0.15697 |
| Full random-effects | **0.15643** | 0.16730 | 0.14145 | 0.14833 | 0.16754 | 0.15621 |
| Hybrid | 0.15652 | 0.16713 | 0.14128 | 0.14839 | 0.16750 | 0.15617 |
| Direct clustering | 0.15726 | 0.16780 | 0.14187 | 0.14864 | 0.16834 | 0.15678 |
| Spherical clustering | 0.15719 | 0.16741 | 0.14198 | 0.14921 | 0.16811 | 0.15678 |
| Autoregressive | 0.15786 | 0.16541 | **0.14028** | 0.14658 | **0.16610** | 0.15525 |
| Hybrid autoregressive | 0.15784 | **0.16515** | 0.14036 | **0.14620** | 0.16613 | **0.15514** |

### 3.3 Goodness-of-fit of hybrid autoregressive model for selected countries

Figure 1 shows the number of weekly cases for the nine countries of exposure with the highest number of reported respiratory illnesses in the period 2015–2019 (China, Hong Kong, India, Indonesia, Nepal, Philippines, Saudi Arabia, Thailand and Vietnam), as well as the corresponding fit of the hybrid autoregressive model. Overall, the mean prediction 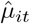 followed the yearly seasonal behavior, with most data lying below the quantiles, indicating a good overall fit, and stable behavior of the weekly cases. Furthermore, the randomized quantile residuals followed the expected uniform distribution, as evidenced by Figure 2. Residual temporal autocorrelations were reduced compared to the raw data (Supplementary Figure S1), though non-negligible temporal autocorrelations persisted for a select number of countries (Nepal, Saudi Arabia, Vietnam). Saudi Arabia represented a notable exception across the three considered diagnostics, indicating that strong peaks during mass gatherings of the annual Hajj could not always be adequately captured by the imposed model structure [19]. Finally, we validated the assumption of independence between countries, as residual spatial correlations were negligible (interquartile range [−0.0419, 0.0478]; Supplementary Figure S2). A similar goodness-of-fit analysis on 2023–2024 data also indicated a satisfactory model fit (see Supplementary Material).

**Fig. 1.**
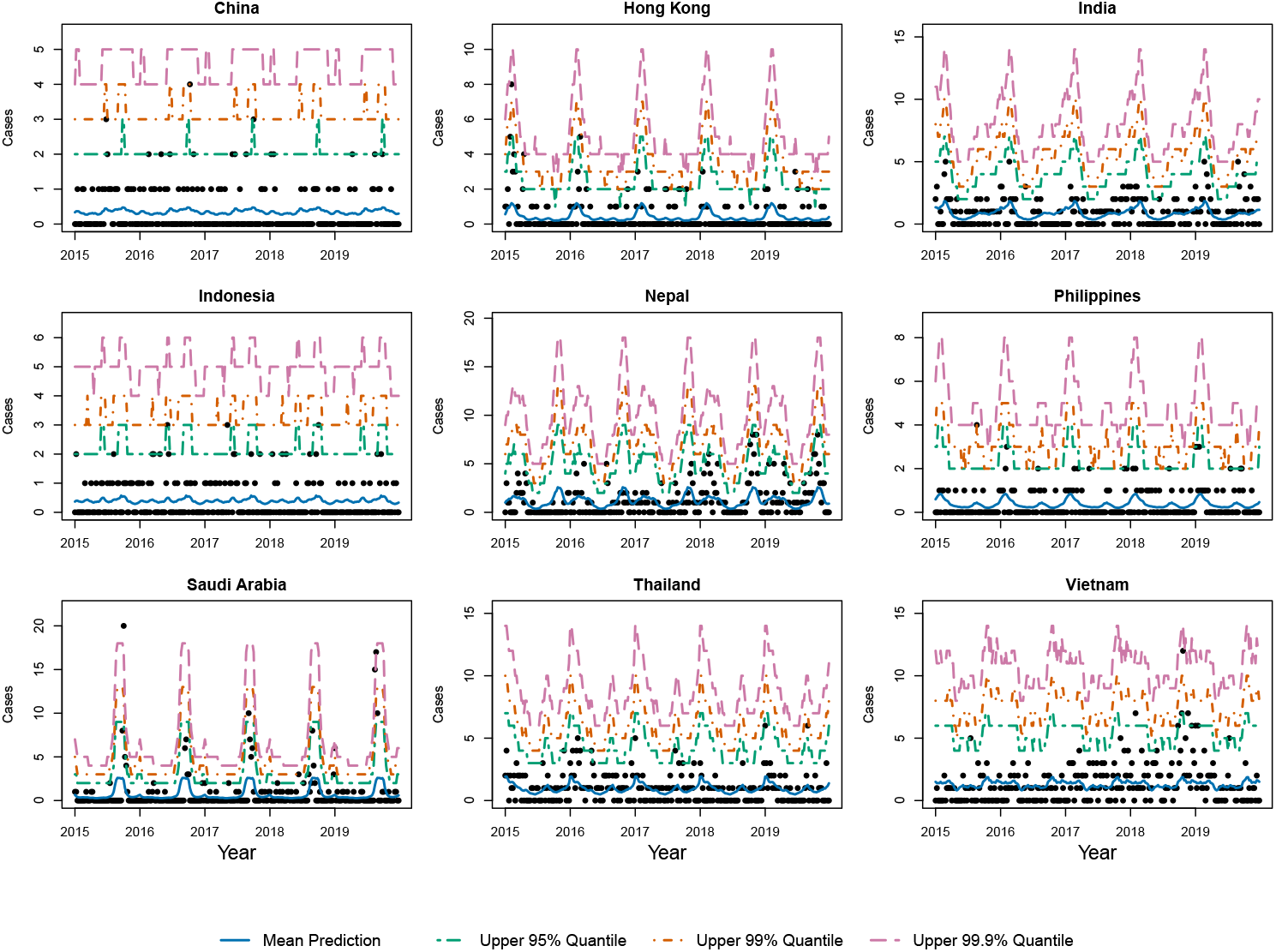
Number of weekly cases for the nine countries with the highest number of overall respiratory illnesses in the period 2015-2019 (China, Hong Kong, India, Indonesia, Nepal, Philippines, Saudi Arabia, Thailand and Vietnam). The fit of the hybrid autoregressive model is visualized by the mean prediction 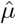 in solid blue, with upper 95%, 99% and 99.9% quantiles indicated by green, orange and purple dotted lines, respectively.

**Fig. 2.**
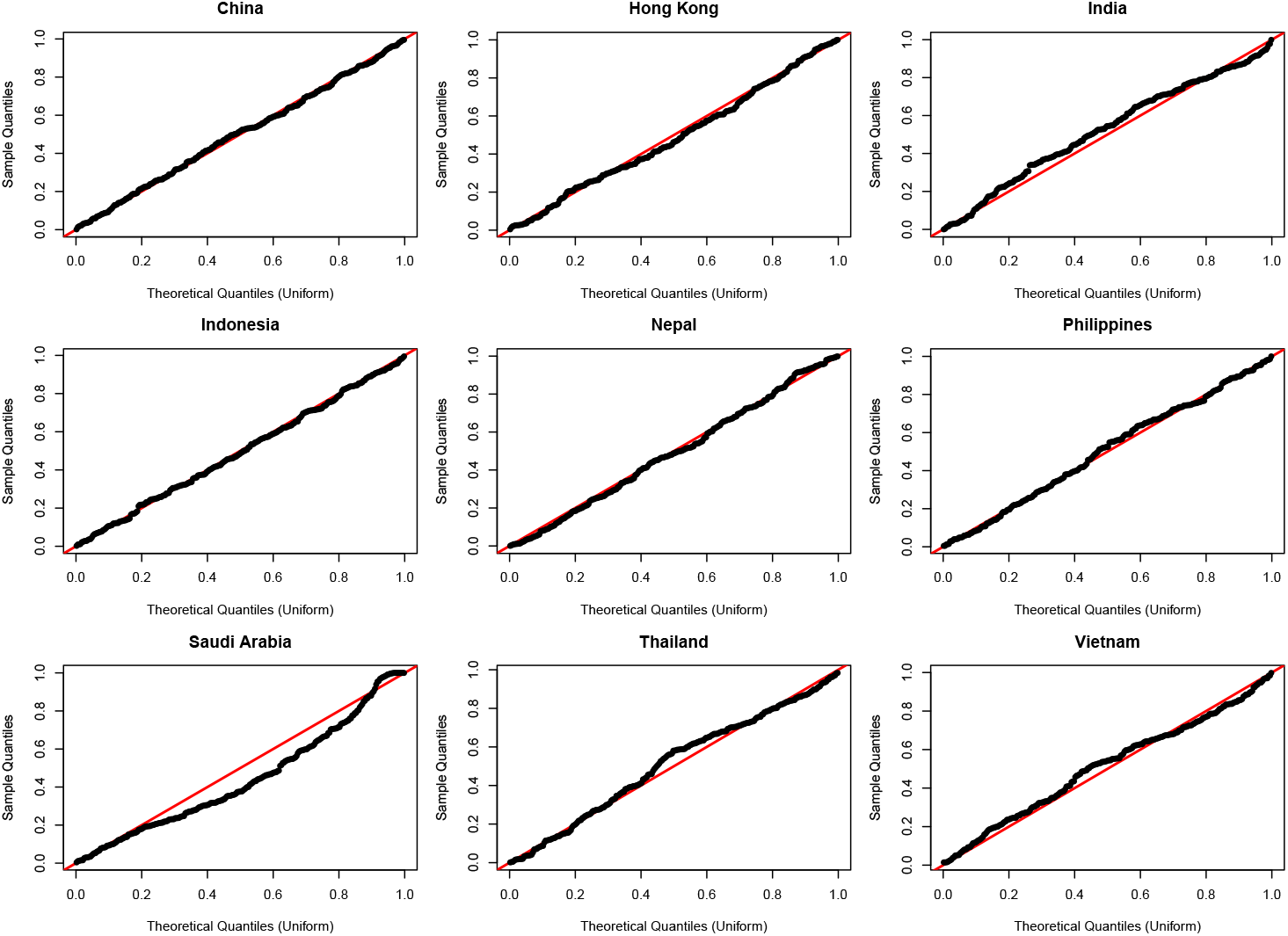
Quantile-Quantile residual plots of the hybrid autoregressive model fitted to data of nine countries with the highest number of overall respiratory illnesses in the period 2015-2019 (China, Hong Kong, India, Indonesia, Nepal, Philippines, Saudi Arabia, Thailand and Vietnam).

### 3.4 Early identification of potential COVID-19 signals in 2020

As shown in Figure 3, signals were raised in China and Italy (out of 64 countries analyzed) in the early weeks of 2020 under the least conservative assumption of no relative increase in travel volume (*c* = 1). In China (54 total cases in 2020, 46.3% potentially novel), a signal occurred under the most conservative assumption on travel volume (*c* = 3) in week 3, though this signal was mainly driven by influenza diagnoses. More notably, a signal occurred under the most conservative assumption (*c* = 3) in week 5 of 2020, driven by respiratory syndromes not attributable to influenza A/B and without a negative COVID-19 test. Italy (41 total cases, 12.2% potentially novel) showed signals in week 8 and 9 under the most conservative assumption (*c* = 3), although these were mostly driven by influenza diagnoses. France (9 total cases, 55.6% potentially novel) and Japan (15 total cases, 60.0% potentially novel) did not trigger any signals, but both came closest to exceeding the least conservative (*c* = 1) threshold among all non-signaling countries, with France in week 4 and Japan in week 5.

**Fig. 3.**
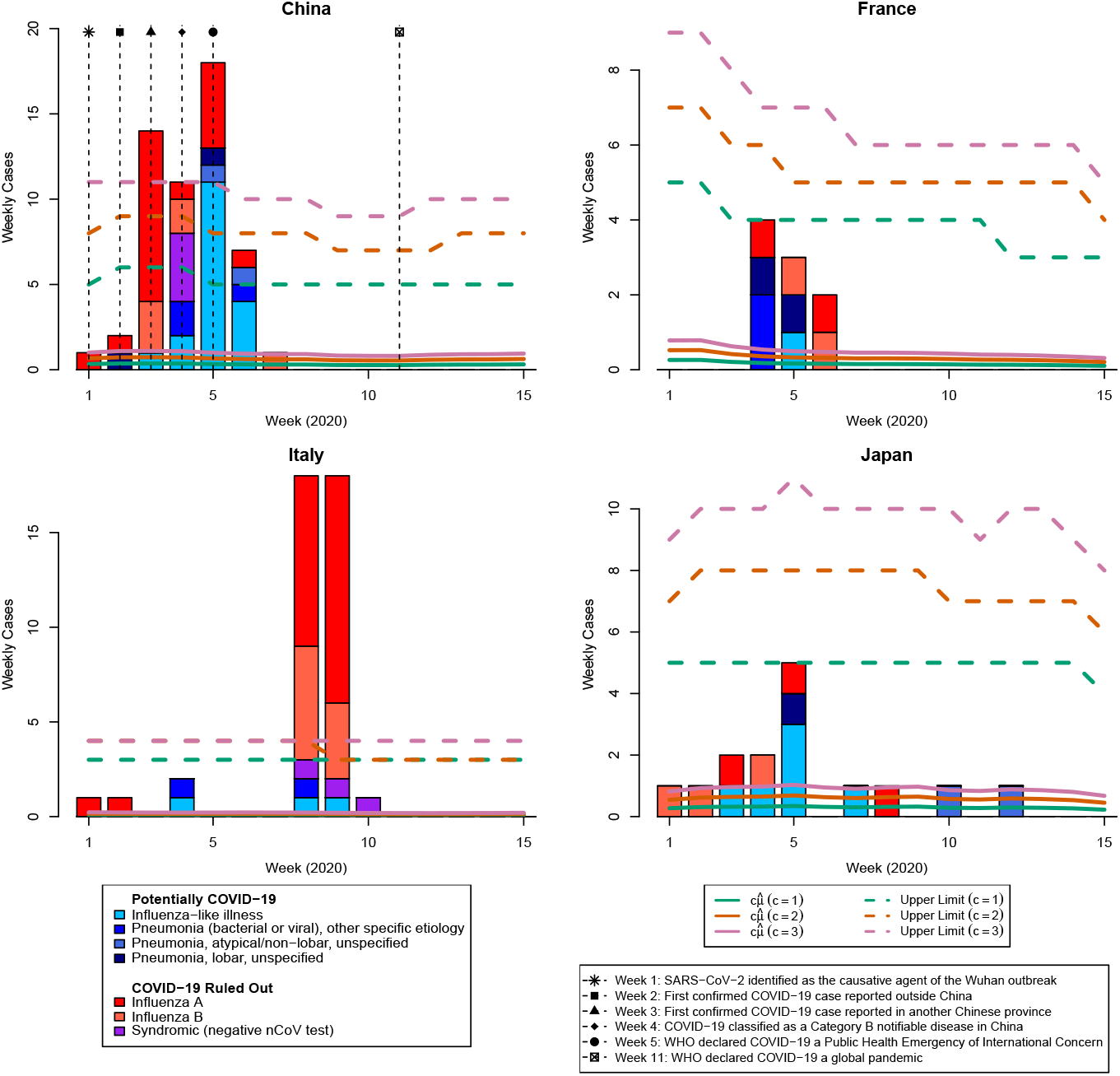
Number of weekly cases for travelers from China, France, Italy, and Japan, respectively, during the initial weeks of 2020. Robust Shewhart charts were set up based on the hybrid autoregressive model fitted to 2015–2019 data and configured with various values for the maximum allowable multiplicative increase in travel volume under non-epidemic conditions (*c* = 1, 2, 3). The global significance level was set so that we expected approximately one false signal every 156 weeks (about three years). We applied a Bonferroni correction and, for 64 countries, set *α* = 1.0016 × 10^−4^, yielding an in-control average run length (ARL_0_) of approximately 10,000 weeks (about 192 years) at the individual country level. Solid lines show scaled mean predictions 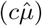, and dotted lines indicate the corresponding upper limits. Out of 64 countries, China and Italy were the only ones that triggered a signal under *c* = 1. France and Japan came close to signaling.

## 4 Discussion

In this work, we demonstrated the potential of traveler surveillance data, combined with an innovative statistical framework, to detect early outbreak signals of respiratory illnesses in the absence of denominators. The study drew on the GeoSentinel network, providing a unique global dataset capturing travel-related illnesses, and used a long observation period (2015–2024) that allowed reliable modeling of epidemiological patterns across pre-pandemic, pandemic, and post-pandemic years. To accurately model baseline behavior, seven GLMMs were extensively compared and the best-performing model was validated for goodness-of-fit, both out-of-sample and in-sample. Robust Shewhart charts were employed to enable outbreak detection, which accounted for unknown denominators by signaling under the assumption of a maximal multiplicative increase in travel volume compared to baseline volume. The practical potential of this approach was illustrated by the successful retrospective identification of an early COVID-19 signal in China using data from the GeoSentinel network.

For context, the COVID-19 outbreak was first recognized in late December 2019, with initial cases associated with the Huanan Seafood Wholesale Market, which included a wet market where wild animals were sold, in Wuhan, China [20, 21]. The outbreak was announced by the Wuhan Health Commission on December 31, 2019, and the causative agent, SARS-CoV-2, was identified by January 7, 2020 [22]. The first case outside of China was identified in a traveler to Thailand on January 13, while a traveler with COVID-19 from Wuhan was identified on January 19 in another Chinese province [20]. COVID-19 was made a class B notifiable disease on January 20, and as SARS-CoV-2 rapidly spread within China and to many other countries, the WHO declared a public health emergency of international concern on January 30, 2020 [22]. At this time, a strong indication of unusual activity among international travelers in the GeoSentinel database returning from China was identified, even under the conservative assumption of *c* = 3 (Figure 3). Based on this week 5 signal, the GeoSentinel network could have leveraged the 20 potential COVID-19 cases identified during weeks 2–5—reported across six sites in four countries (France, Germany, Japan, and the United States)—to alert the respective public health authorities about the risk of COVID-19 importations, well before the WHO declared COVID-19 a global pandemic on March 11, 2020 [23].

Although Figure 3 showed that no early actionable signals were detected for Italy, Japan, or France, these countries revealed important limitations of the approach. Namely, Italy experienced an exponential surge in COVID-19 cases during the early pandemic, severely straining its healthcare system [24, 25], although the number of reported potential COVID-19 cases to the GeoSentinel database remained limited. While the epidemiological interactions between COVID-19 and influenza epidemics are complex (see [26] for further discussion), we hypothesize that increased public awareness and expanded diagnostic testing following this surge inflated reported influenza A and B cases, producing the week 8–9 signals. Japan, which was third to report a COVID-19 case after China and Thailand [27], did not trigger an alert, and France, despite confirming Europe’s first cases on January 24, 2020 [24], remained below the *c* = 1 threshold. Other early-affected countries, such as Thailand and the USA [21, 27], also showed no signals, likely reflecting early reduction in travel to these areas or behavioral factors such as a reduced likelihood of seeking care at travel clinics in favor of urgent care or primary care for acute lower respiratory illnesses.

Despite the strong surge in cases in China, the GeoSentinel network did not recognize the nature of the outbreak, partly due to the absence of baseline epidemiology for syndromic diagnoses across global regions and the lack of reliable travel volume data. The dynamic and often unknown nature of travel volume complicates baseline modeling, as it limits the use of climatological or geographically driven variability typically employed in respiratory illness surveillance. For instance, countries in the Northern Hemisphere generally exhibit peak respiratory illness activity during November–February, while those in the Southern Hemisphere peak during May–September [28]. However, latent and fluctuating travel volumes confound these seasonal patterns in traveler-based surveillance. This challenge was exemplified by surges in cases in Saudi Arabia during the annual Hajj mass gathering [19] (see Figure 1). Therefore, more flexible, data-driven approaches were considered in this work. For modeling counts in multiple geographic regions or areas, mixed-effects models with lognormally distributed random intercepts are often the primary choice [12]. These models are advantageous because they limit the number of parameters when regional heterogeneity is expected. However, in this context, the conventional assumption of lognormally distributed random intercepts may not adequately capture the heavy-tailed distribution of international travel volumes, where a small subset of destinations attracts the majority of visitors [29]. Our comparative analysis reflected this, as the full random-effects model was outperformed by the hybrid model. Furthermore, incorporating latent temporal autocorrelation substantially improved model fit and reduced residual spatial-temporal correlations while maintaining the assumption of constant year-to-year dynamics. The high performance of the autoregressive models suggested that these terms played a central role in capturing the short-term serial dependencies inherent in infectious disease data. The hybrid autoregressive model outperformed the autoregressive model, which might be because many countries in the dataset contained sparse observations, for which stronger parametric assumptions could assist with more stable model estimation. Our goodness-of-fit analysis showed that some years exhibit notably higher seasonal peaks, a phenomenon commonly documented in influenza literature [28]. While incorporating limited year-to-year variability might improve model performance, its marginal effect and the risk of masking unusually high peaks that could signal emerging threats led us to adopt a simpler model that did not explicitly account for year-to-year variation.

The GeoSentinel data presented a few limitations. As with many studies in this field (e.g., [28]), a key limitation is that GeoSentinel data may not capture sub-national variability, while countries with regional differences may require more localized modeling approaches. While our proof-of-principle deliberately excludes COVID-19 diagnostic codes, relying solely on syndromic LRTI case increases as would be available in real-time surveillance, our analysis does not account for reporting delays, which would have caused alarms to be rung too late for meaningful public health intervention had the framework been deployed in real time at the onset of the pandemic. Reducing reporting delays is currently a major priority for GeoSentinel, and statistical approaches that explicitly model these lags to improve nowcasts (e.g. [30, 31]) warrant serious consideration.

Also the presented methodology has several limitations worth considering. The initial presentation of a novel pathogen, including its specific syndromic or etiologic features, may be unknown during the early stages of an outbreak. To address this, multiple respiratory diagnoses were aggregated into a single analysis group. Aggregating diagnoses increases the chance of detecting a signal spread thinly across several individual codes, compared with monitoring each code in isolation. However, it may also inflate baseline estimates and slightly reduce sensitivity, as cases unrelated to the pathogen of interest confound the signal. To mitigate this, after a signal induced by aggregated case counts, we contrasted potential COVID-19 case counts against the aggregated baseline.

The risk of inflating baseline estimates favors restricting the analysis to clearly relevant codes, but doing so risks discarding less obviously relevant codes whose evidential value may nonetheless be substantial, a known challenge in syndromic surveillance [9]. Our post-hoc examination illustrated this phenomenon: a generic viral syndrome code, excluded from the primary analysis for its non-specific definition, showed a major increase in Western Europe, particularly Italy, during the early weeks of 2020, consistent in timing and geography with the emergence of COVID-19. Going forward, surveillance networks should prioritize two fronts. At the data-entry level, this means refining how case definitions balance specificity against generality, supported by consistent coding across sites. At the methodological level, it means developing detection methods which are more robust to code misclassification, for instance by casting a wider net of candidate codes and weighting their contributions by their evolving evidential value during an emerging outbreak [32].

In this work, conservative design parameters (ARL_0_ and *c*) were chosen to strengthen the proof-of-principle that the framework can detect outbreaks of high public health importance. However, operational deployment would benefit from further optimization of these parameters, with the optimal trade-off between false positives and false negatives depending on the relative costs of each: including available resources for investigating false alarms (false positives), the epidemiological context, and the consequence of missing an emerging outbreak (false negatives). Still, the proposed system serves as an early warning tool rather than definitive evidence of an outbreak, and its successful implementation will rely on continued collaboration between data scientists and public health officials to interpret signals and validate underlying assumptions.

## 5 Conclusion

This study focused on understanding and enhancing the utility of international traveler data for surveillance of respiratory illnesses with pandemic potential, specifically from the GeoSentinel network. We compared several generalized linear mixed models and validated the preferred hybrid autoregressive model based on its goodness-of-fit to establish a suitable model for baseline epidemiology. Although reliable denominator data are unavailable, we described an inferential framework using robust Shewhart charts that enables outbreak detection under conservative assumptions about maximum increases in travel volume. We demonstrated the framework’s practical applicability by the early detection of an outbreak of unknown etiology — later identified as COVID-19 — through an increase in syndromic acute LRTI cases.

The respiratory syndromic signal in China in early 2020, detected under the conservative assumption of up to a threefold increase in travel volume and against a validated 2015-2019 baseline model, shows that the network held the potential to contribute to COVID-19 knowledge in the early stages of the pandemic by sharing international traveler data with political leaders and health authorities. Together with the satisfactory model fit for post-pandemic data, this suggests that GeoSentinel could contribute to global health security by providing early sentinel surveillance signals for respiratory illness outbreaks.

## Availability of data and materials

Aggregate epidemiological and clinical data supporting the findings of this study are included in the article. De-identified individual-level data are not publicly available because they were collected through routine public health surveillance and are subject to data protection and institutional restrictions. R functions used for statistical modeling can be found at https://github.com/SHeidema/ilitools.

## Acknowledgements

We acknowledge the GeoSentinel Surveillance Network and the GeoSentinel Foundation, Inc. for their support in data collection, coordination and infrastructure enabling this collaborative analysis.

## Note

The findings and conclusions in this report are those of the authors and do not necessarily represent the official position of the Centers for Disease Control and Prevention.

## Ethical statement

This study used routine public health surveillance data from the GeoSentinel Surveillance Network, involving no intervention beyond standard clinical care. This activity was reviewed by the Centers for Disease Control and Prevention (CDC), deemed not research, and conducted consistent with applicable federal law and CDC policy.^1^ Institutional review board approval was therefore not required.

## Appendix A Supplementary Methods: Alternative Models for Baseline Epidemiology

In this work, it is assumed that

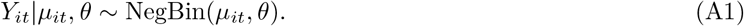

Several structures for *µ*_*it*_ were considered. The hybrid autoregressive model is used throughout this work. The other model structures used for comparison are described here. As the counts are often small, two approaches that cluster the estimated parameters of the full fixed effects model (A2) were considered.

### Full fixed-effects model

The full fixed-effects model is structured as:

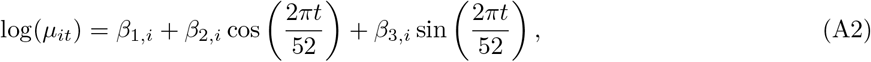

where *β*_1,*i*_, *β*_2,*i*_ and *β*_3,*i*_ are fixed parameters for country *i*.

### Full random-effects model

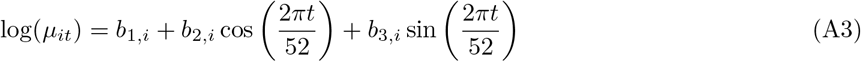

Here, the random country-specific coefficients *b*_1,*i*_, *b*_2,*i*_, and *b*_3,*i*_ are assumed to follow independent normal distributions:

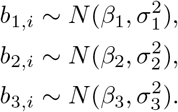

### Hybrid model

The hybrid model employs a fixed effect for the intercept, but random effects for seasonal effects. Specifically, it is defined as:

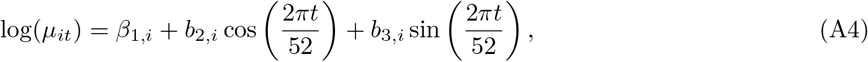

where the seasonal components’ coefficients, *b*_2,*i*_ and *b*_3,*i*_, follow independent normal distributions.

### Direct clustering

Here, we fit the full fixed effects model (A2), and cluster the seasonal coefficients 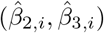 using *k*-means in ℝ^2^. The number of clusters is determined using cross-validation. The cluster of country *i* is denoted as *j*. Then the model is refitted using less seasonal components:

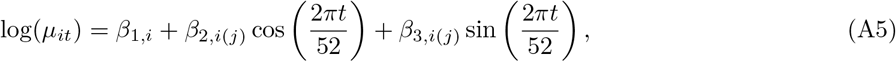

### Spherical clustering

Feature-based approaches reduce time series to a set of summary statistics or model parameters, which are then clustered using standard clustering algorithms to identify patterns or groups. Specifically, we start by obtaining country-specific baseline 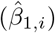, amplitude 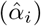 and phase 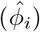 estimates for all *i*, as well as an estimate 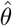 for the dispersion parameter. Instead of fitting a series-specific nonlinear model (i.e., the seasonal structure in (A10) with *J* = *I*), note that we can also write the mean of the country-specific model as

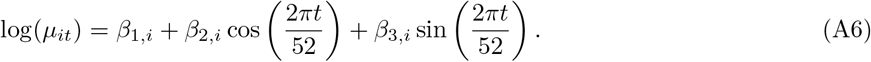

In the spherical approach, we note that

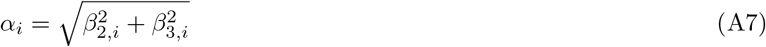

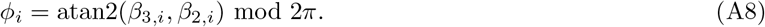

Clustering countries with similar phase parameters when the corresponding amplitudes are not practically relevant, as done in direct clustering, might be suboptimal. Therefore, we classify series based on whether they exhibit seasonal behavior or not. Using approximate bivariate normality of 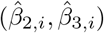, we apply a Hotellings *T* ^2^ test to test ℋ_0,*i*_ : *α*_*i*_ = 0 at significance level *γ*. If ℋ_0,*i*_ is not rejected, country *i* is assigned to a non-seasonal cluster. The phase estimates 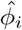 for which ℋ_0,*i*_ is rejected at significance level *γ* are clustered according to spherical *k*-means, aiming to maximize the cosine similarity measure

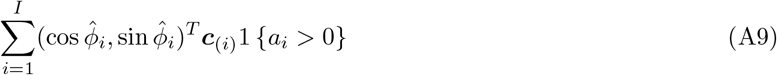

between the sample 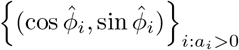 and *k* centroids ***c***_1_, …, ***c***_*k*_ ∈ S^1^, where ***c***_(*i*)_ represents the centroid of the cluster containing country *i* [33]. The significance level *γ*, and the number of clusters, are optimized using cross-validation. Given the newly induced structure, we refit the model as:

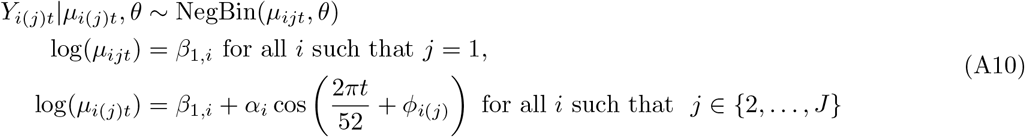

### Autoregressive

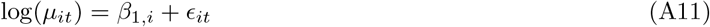

The term *ϵ*_*it*_ is normally distributed, and reflects a periodic temporal autocorrelation structure to account for the annual cycle:

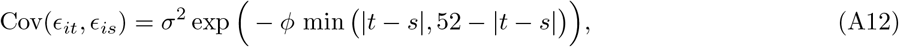

where *t* and *s* are week indices, and observations from different countries of exposure remain uncorrelated (Cov(*ϵ*_*it*_, *ϵ*_*js*_) = 0 if *i* ≠ *j*).

## Appendix B Supplementary Results: Goodness-of-Fit (Pre-Pandemic)

**Fig. B1: Supplementary Figure S1.**
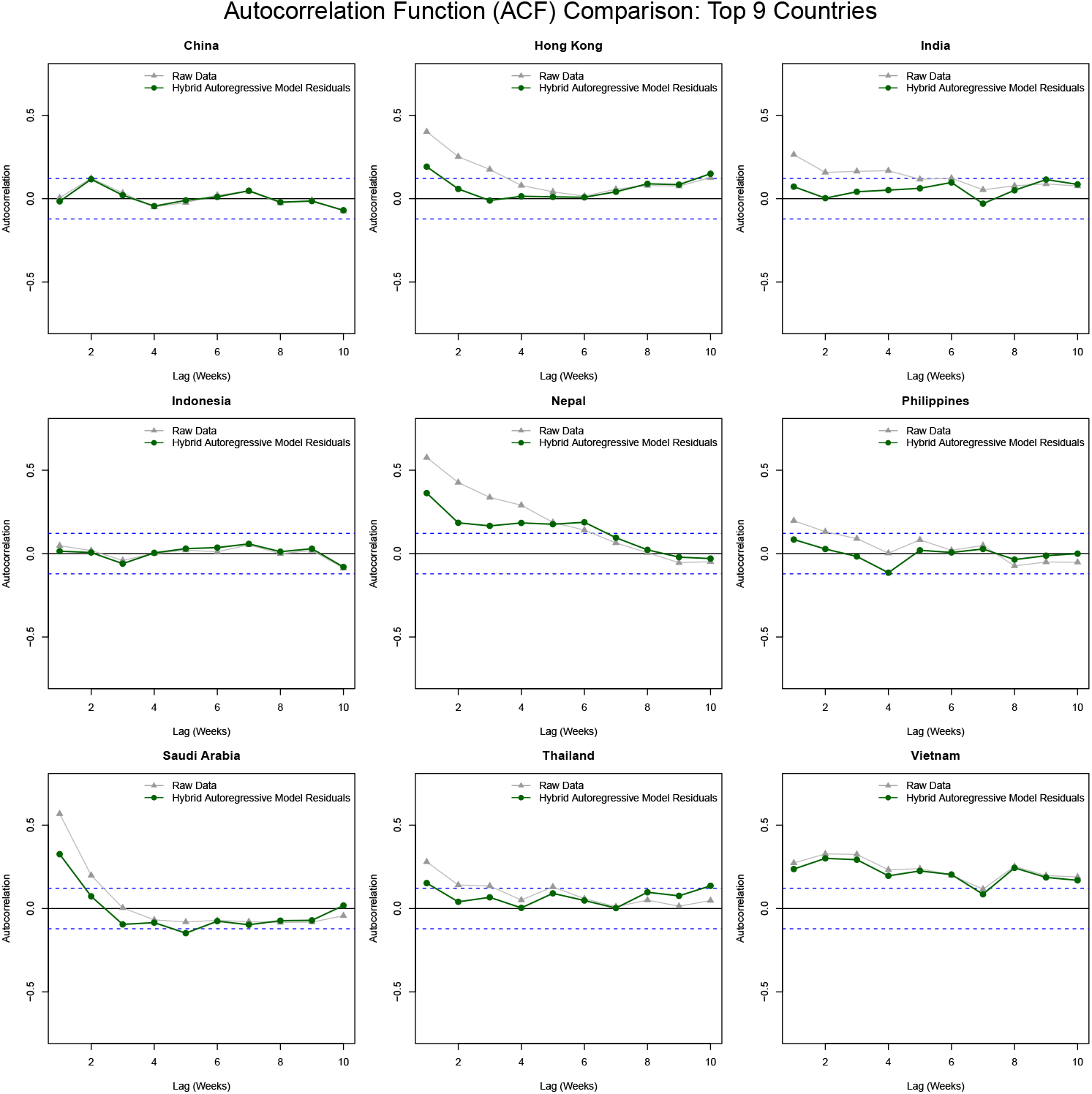
Residual temporal autocorrelation for lags 1-10 of the hybrid autoregressive model (green) fitted to data of the nine countries with the highest number of reported respiratory illnesses in 2015-2019 (China, Hong Kong, India, Indonesia, Nepal, Philippines, Saudi Arabia, Thailand, and Vietnam), compared to the raw data ACF (grey). Blue dotted lines indicate 95% significance bounds 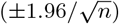.

**Fig. B2: Supplementary Figure S2.**
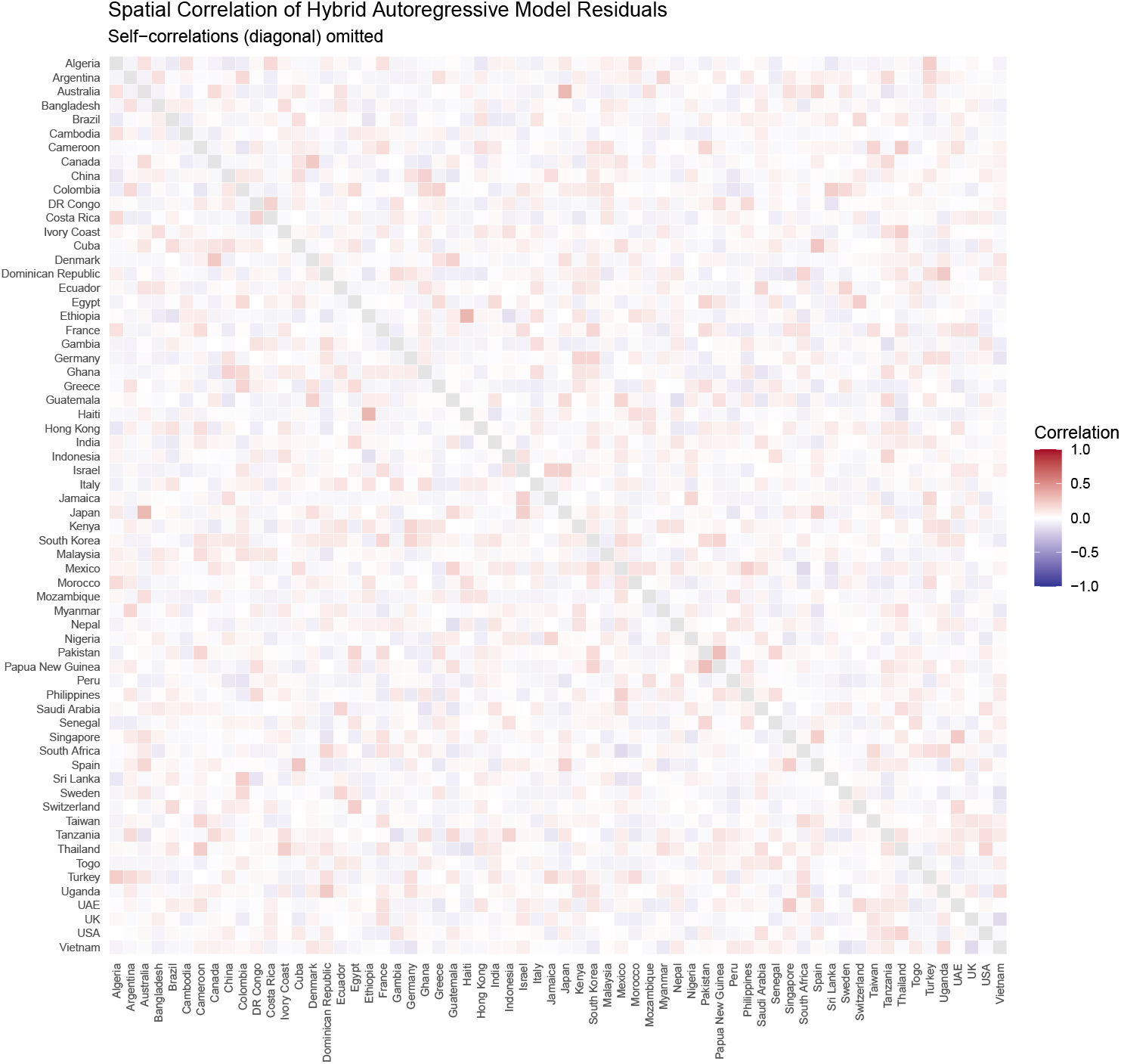
Residual spatial autocorrelation of the hybrid autoregressive model fitted to 2015-2019 data of all 64 countries.

## Appendix C Supplementary Results: Goodness-of-Fit (Post-Pandemic)

Figure C3 shows the fit of the hybrid autoregressive model for the nine countries with the highest number of reported respiratory illnesses in 2023–2024. The mean prediction 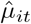 captured the overall seasonal dynamics well, with observed data generally falling within the upper quantile bounds. The randomized quantile residuals followed the expected uniform distribution (Figure C4), indicating satisfactory model performance. Notably, model fit improved for Saudi Arabia compared with the pre-pandemic period, showing a more stable post-pandemic pattern in weekly case counts.

**Fig. C3: Supplementary Figure S3.**
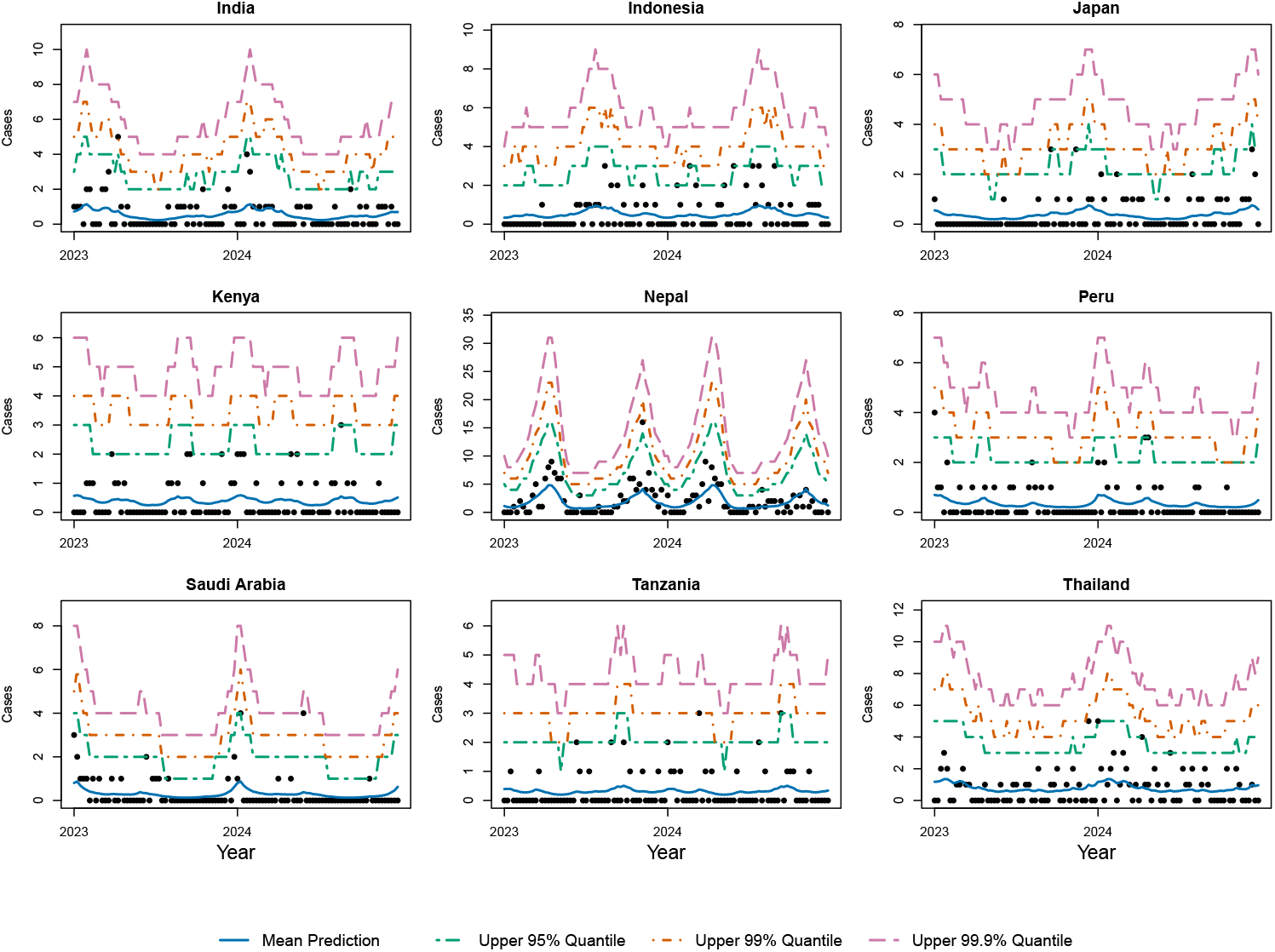
Number of weekly cases for the nine countries with the highest number of overall respiratory illnesses in the period 2023-2024 (India, Indonesia, Japan, Kenya, Nepal, Peru, Saudi Arabia, Tanzania and Thailand). The fit of the hybrid autoregressive model is visualized by the mean prediction 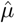 in solid blue, with upper 95%, 99% and 99.9% quantiles indicated by green, orange and purple dotted lines, respectively.

**Fig. C4: Supplementary Figure S4.**
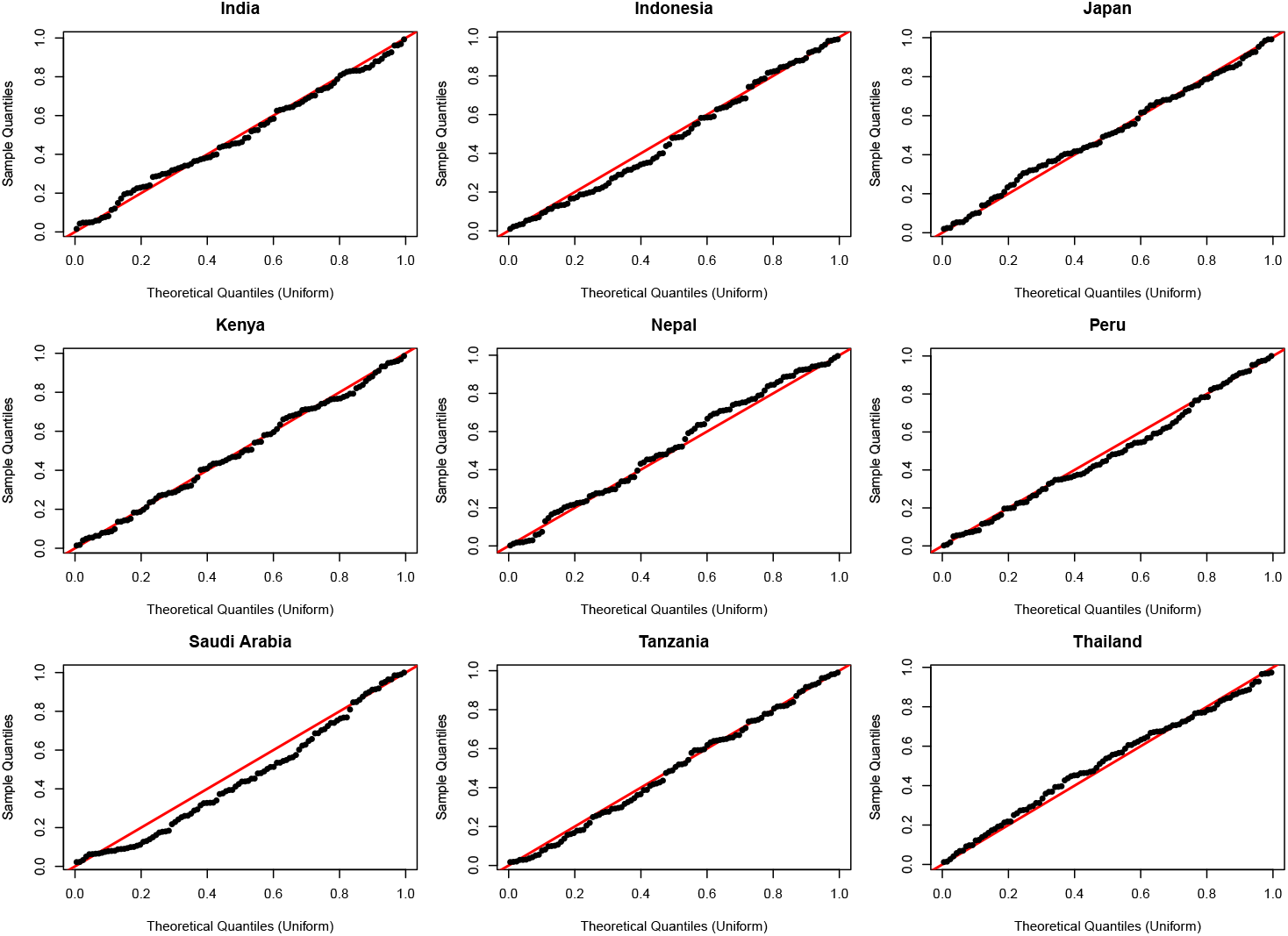
Quantile-Quantile residual plots of the hybrid autoregressive model fitted to data of nine countries with the highest number of overall respiratory illnesses in the period 2023-2024 (India, Indonesia, Japan, Kenya, Nepal, Peru, Saudi Arabia, Tanzania and Thailand).

**Fig. C5: Supplementary Figure S5.**
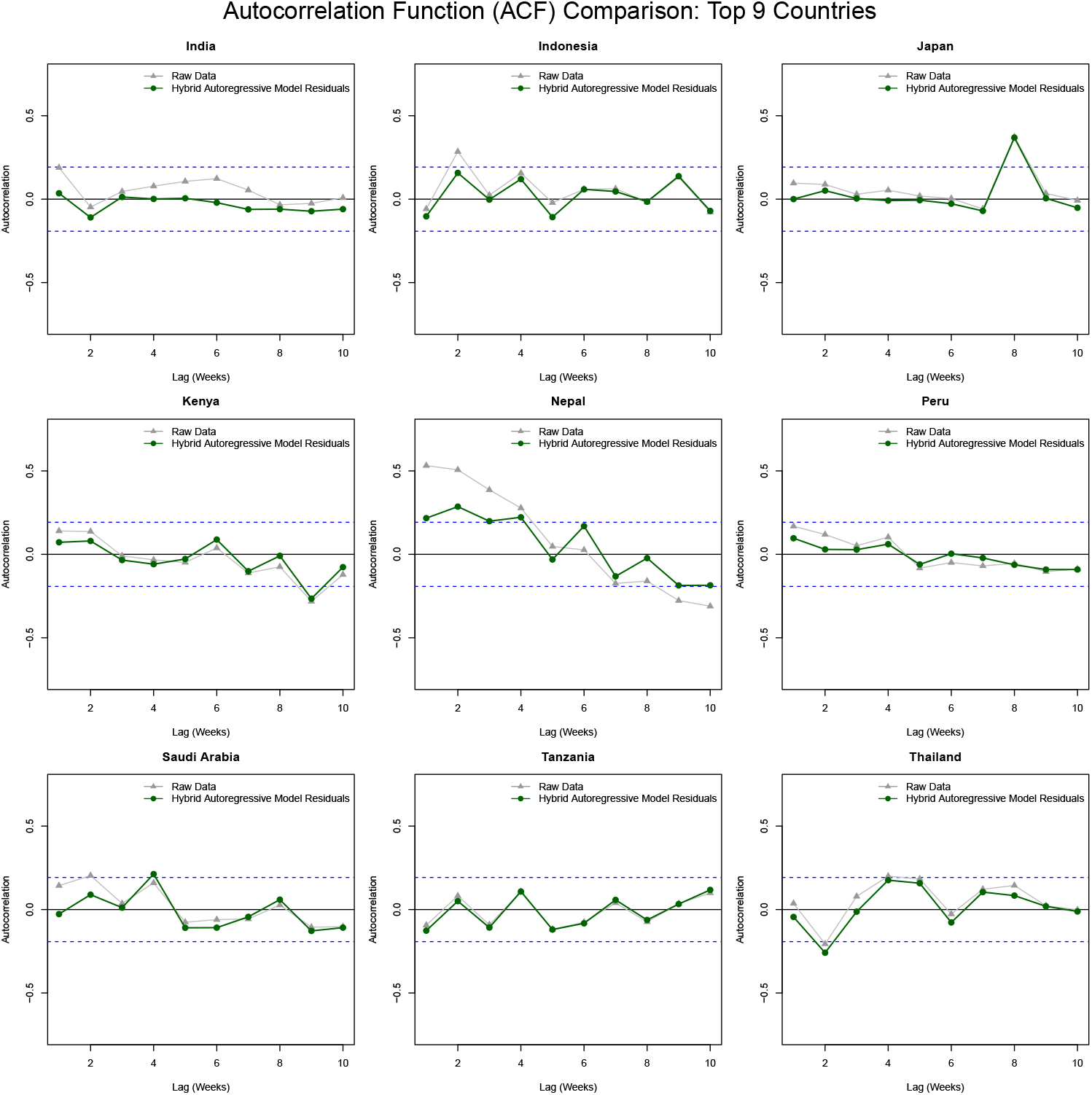
Residual temporal autocorrelation for lags 1-10 of the hybrid autoregressive model (green) fitted to data of the nine countries with the highest number of reported respiratory illnesses in 2023-2024 (India, Indonesia, Japan, Kenya, Nepal, Peru, Saudi Arabia, Tanzania and Thailand), compared to the raw data ACF (grey). Blue dotted lines indicate 95% significance bounds 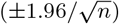.

**Fig. C6: Supplementary Figure S6.**
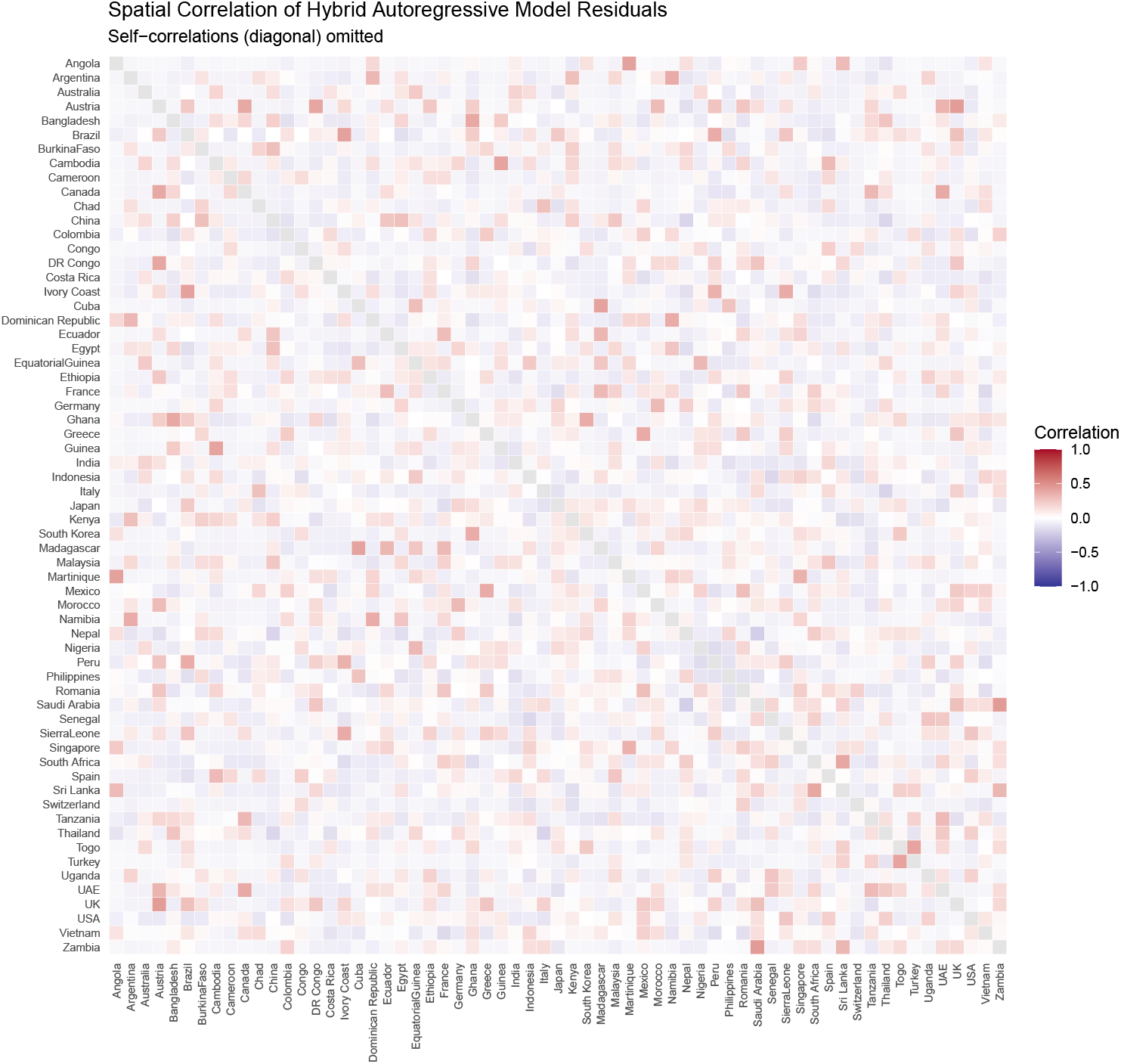
Residual spatial autocorrelation of the hybrid autoregressive model fitted to 2023-2024 data of all 63 countries.

## Appendix D Supplementary Results: Sensitivity Analysis

We conducted a sensitivity analysis to assess the impact of including conditions with uncertain classification (acute bronchitis, ARDS, and viral respiratory infection of other specific etiology) on overall findings.

Table D1 summarizes the number of reported cases across the study periods given the inclusion of these conditions with uncertain classification. Including these conditions resulted in a modest increase in total records (*n* = 4, 567 for 2015–2019, *n* = 355 for 2020, and *n* = 1, 404 for 2023–2024) and a slightly broader geographic representation (68 and 67 countries, respectively). The proportional distribution of etiologic and syndromic diagnoses remained similar, with influenza A and influenza-like illness continuing to represent the leading etiologic and syndromic categories. The inclusion of acute bronchitis accounted for most of the additional cases, while new entries for viral respiratory infection of other specific etiology and ARDS contributed minimally.

The goodness-of-fit assessment (Figures D7 and D8) confirmed that model performance remained robust when including conditions with uncertain classification for the period 2015–2019. As in the main analysis, the hybrid autoregressive model adequately captured the seasonal patterns of weekly case counts across the nine countries with the highest case volumes, with the mean predictions 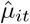 closely tracking observed data. The randomized quantile residuals continued to approximate a uniform distribution, supporting model adequacy. Deviations persisted for Saudi Arabia, where strong peaks during the Hajj period remained only partially captured by the model structure.

As shown in Figure D11, inclusion of conditions with uncertain classification did not change the overall signal detection results. Signals were again observed in China and Italy during the early weeks of 2020 under *c* = 3, with comparable magnitudes to the main analysis. No additional countries reached the signaling threshold, and France and Japan remained the closest non-signaling locations.

The goodness-of-fit assessment for the post-pandemic period (Figures D12 and D13) confirmed that the model performance remained robust when including conditions with uncertain classification. The mean prediction 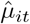 continued to capture the seasonal behavior of weekly case counts across the nine countries with the highest number of reported illnesses, with most observations lying within the modeled quantile bounds. The randomized quantile residuals maintained an approximately uniform distribution, indicating an adequate fit. As in the main analysis, model performance improved for Saudi Arabia relative to the pre-pandemic period, reflecting a more stable post-pandemic trend.

**Table D1: Supplementary Table S1.** Reported acute lower respiratory illness cases by condition and inclusion status, across 68 countries (2015–2019), 68 countries (2020), and 67 countries (2023–2024), including conditions with differing expert classification as acute LRTIs. Countries with fewer than 2 average annual acute LRTI cases were excluded.

| Included Conditions |  |  |  |  |  |
| --- | --- | --- | --- | --- | --- |
| 2015–2019 (68 countries) | *n* | 2020 (68 countries) | *n* | 2023–2024 (67 countries) | *n* |
| *Etiologic* |  |  |  |  |  |
| Influenza A | 836 | Influenza A | 2 | COVID-19 | 423 |
| Influenza B | 358 | Influenza B | 102 | Influenza A | 290 |
| Legionnaires' Disease | 45 | Legionnaires' Disease | 37 | Influenza B | 94 |
| Pneumonia (bacterial or viral, other specific etiology) | 274 | Pneumonia (bacterial or viral, other specific etiology) | 21 | Pneumonia (bacterial or viral, other specific etiology) | 69 |
|  |  |  |  | Viral respiratory infection, other specific etiology | 40 |
| *Syndromic* |  |  |  |  |  |
| Acute Bronchitis | 1055 | Acute Bronchitis | 70 | Acute Bronchitis | 298 |
| Influenza-like illness | 1247 | Influenza-like illness | 78 | Influenza-like illness | 137 |
| Pneumonia, atypical/non-lobar, unspecified | 259 | Pneumonia, atypical/non-lobar, unspecified | 23 | Pneumonia, atypical/non-lobar, unspecified | 43 |
| Pneumonia, Lobar, Unspecified | 493 | Pneumonia, Lobar, Unspecified | 22 | Pneumonia, Lobar, Unspecified | 110 |
| **Total (Included)** | **4567** | **Total (Included)** | **355** | **Total (Included)** | **1404** |
| Excluded Conditions |  |  |  |  |  |
| *Etiologic* |  |  |  |  |  |
| COVID-19 | 0 | COVID-19 | 645 | Legionnaires' Disease | 3 |
| Influenza, Avian | 0 | Influenza, Avian | 0 | Influenza, Avian | 0 |
| MERS-Co-V | 0 | MERS-Co-V | 0 | MERS-Co-V | 0 |
| SARS | 0 | SARS | 0 | SARS | 0 |
| Viral respiratory infection, other specific etiology | 1 | Viral respiratory infection, other specific etiology | 4 |  |  |
| *Syndromic* |  |  |  |  |  |
| Acute respiratory distress syndrome (ARDS) | 3 | Acute respiratory distress syndrome (ARDS) | 2 | Acute respiratory distress syndrome (ARDS) | 4 |
| Pneumonia, pneumococcal | 9 | Pneumonia, pneumococcal | 3 | Pneumonia, pneumococcal | 9 |
| **Total (Excluded)** | **13** | **Total (Excluded)** | **654** | **Total (Excluded)** | **16** |
| **Total** | **4580** | **Total** | **1009** | **Total** | **1420** |

### D.1 Sensitivity analysis: Goodness-of-Fit (pre-pandemic)

**Fig. D7: Supplementary Figure S7.**
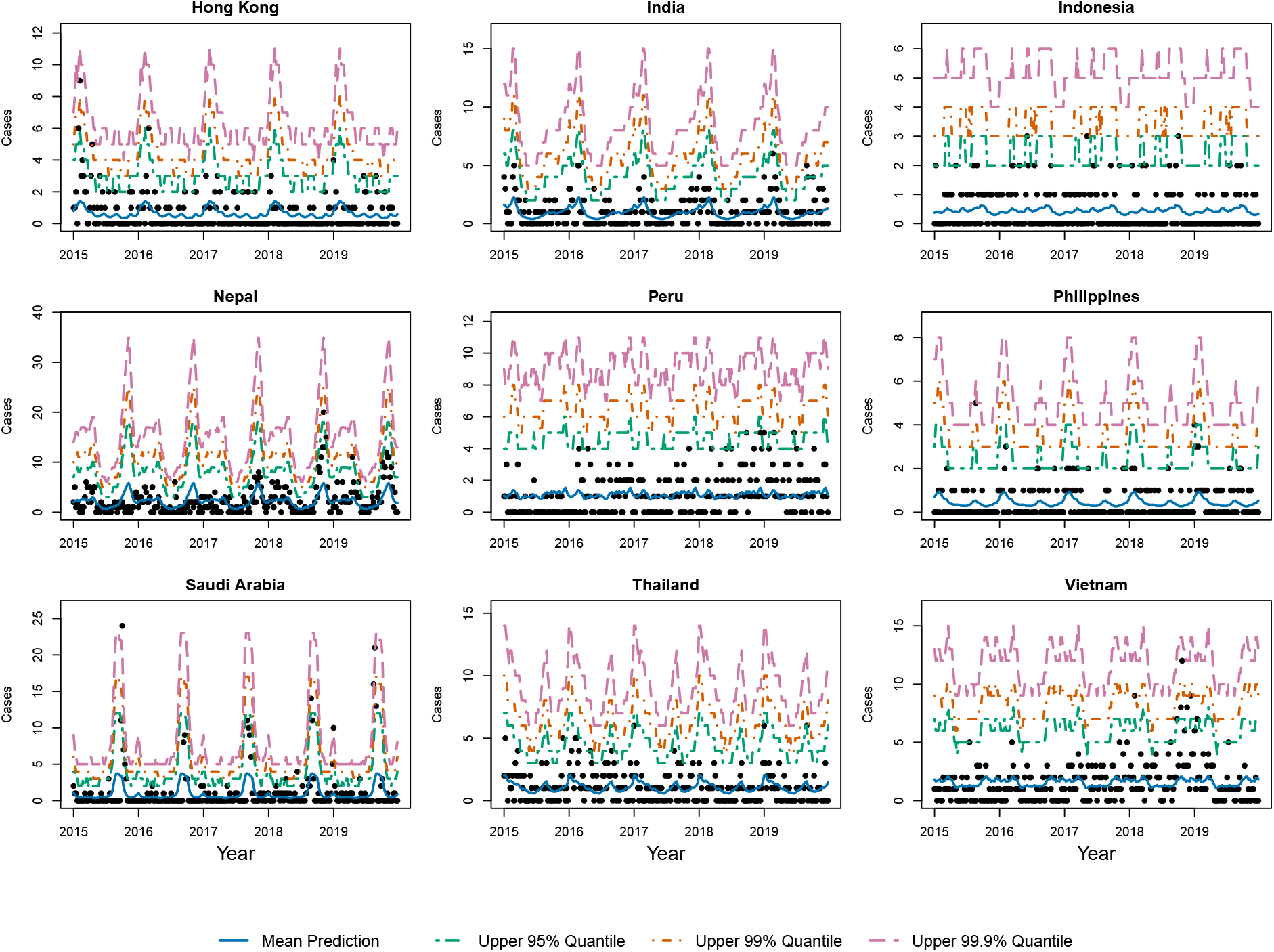
Number of weekly cases for the nine countries with the highest number of overall respiratory illnesses in the period 2015-2019 (Hong Kong, India, Indonesia, Nepal, Peru, Philippines, Saudi Arabia, Thailand and Vietnam), including conditions with differing expert classification as acute LRTIs. The fit of the hybrid autoregressive model is visualized by the mean prediction 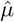 in solid blue, with upper 95%, 99% and 99.9% quantiles indicated by green, orange and purple dotted lines, respectively.

**Fig. D8: Supplementary Figure S8.**
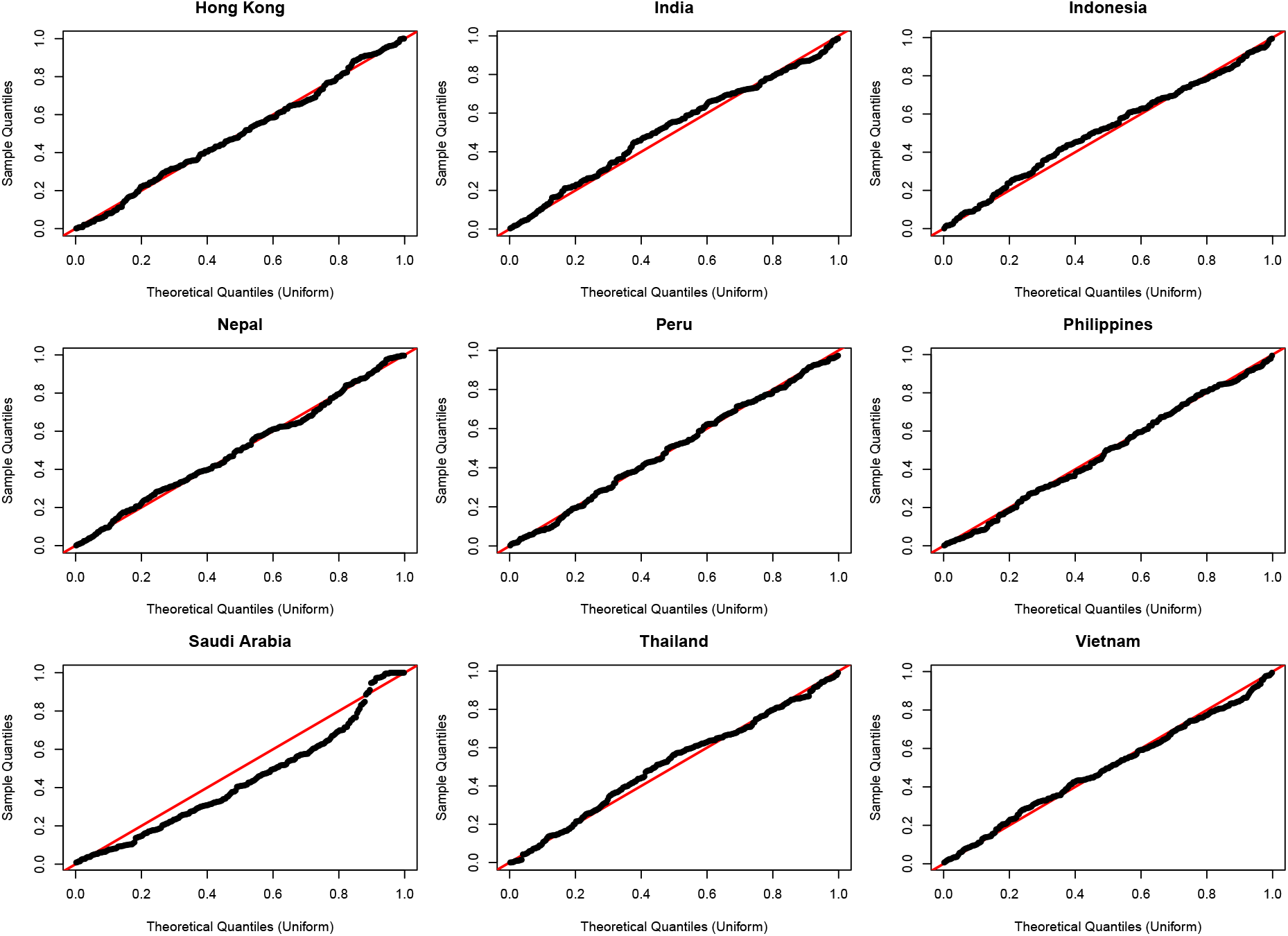
Quantile-Quantile residual plots of the hybrid autoregressive model fitted to data of nine countries with the highest number of overall respiratory illnesses in the period 2015-2019 (Hong Kong, India, Indonesia, Nepal, Peru, Philippines, Saudi Arabia, Thailand and Vietnam), including conditions with differing expert classification as acute LRTIs.

**Fig. D9: Supplementary Figure S9.**
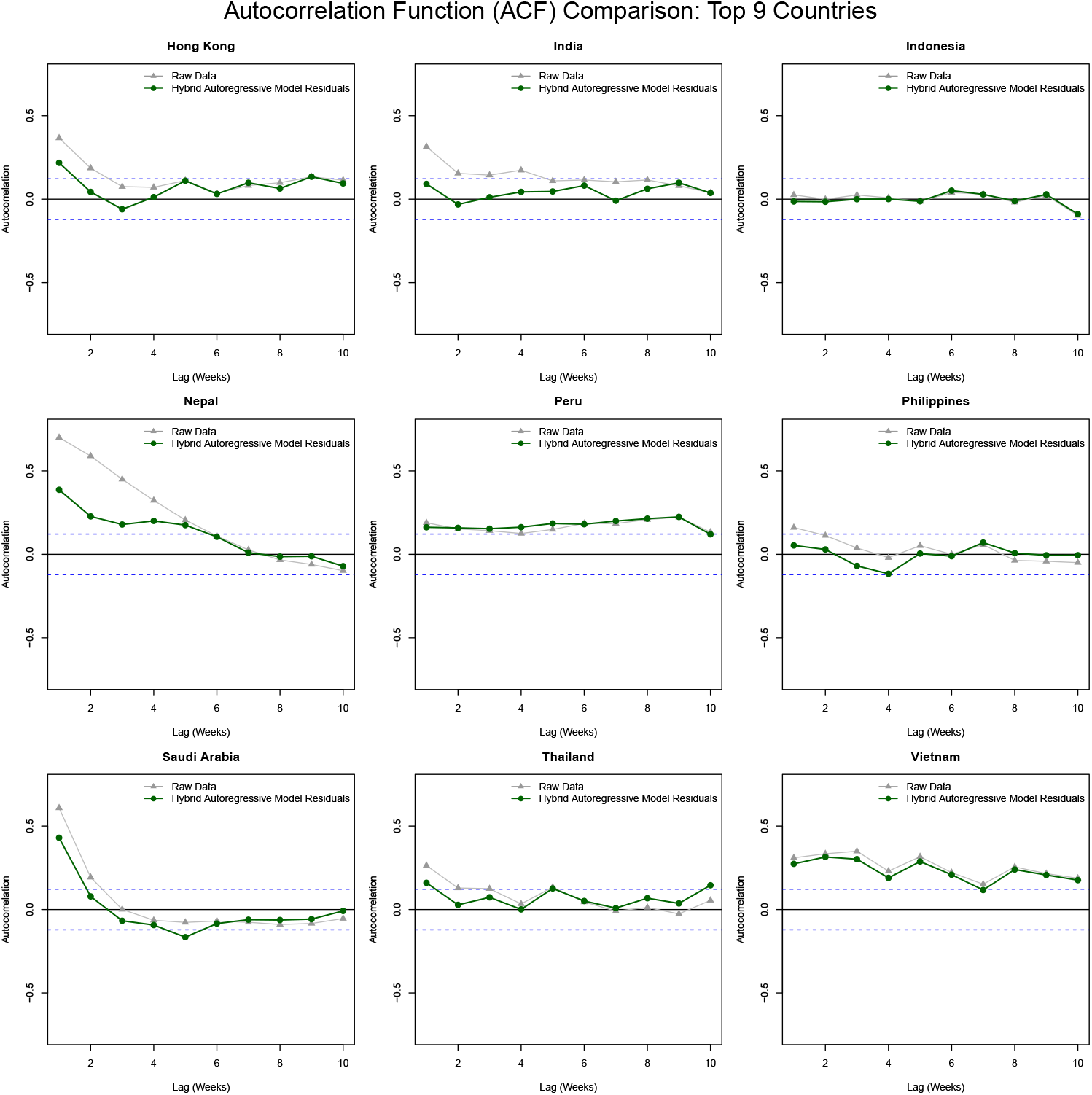
Residual temporal autocorrelation for lags 1-10 of the hybrid autoregressive model (green) fitted to data of the nine countries with the highest number of reported respiratory illnesses (including conditions with differing expert classification as acute LRTIs) in 2015-2019 (Hong Kong, India, Indonesia, Nepal, Peru, Philippines, Saudi Arabia, Thailand and Vietnam), compared to the raw data ACF (grey). Blue dotted lines indicate 95% significance bounds 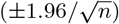.

**Fig. D10: Supplementary Figure S10.**
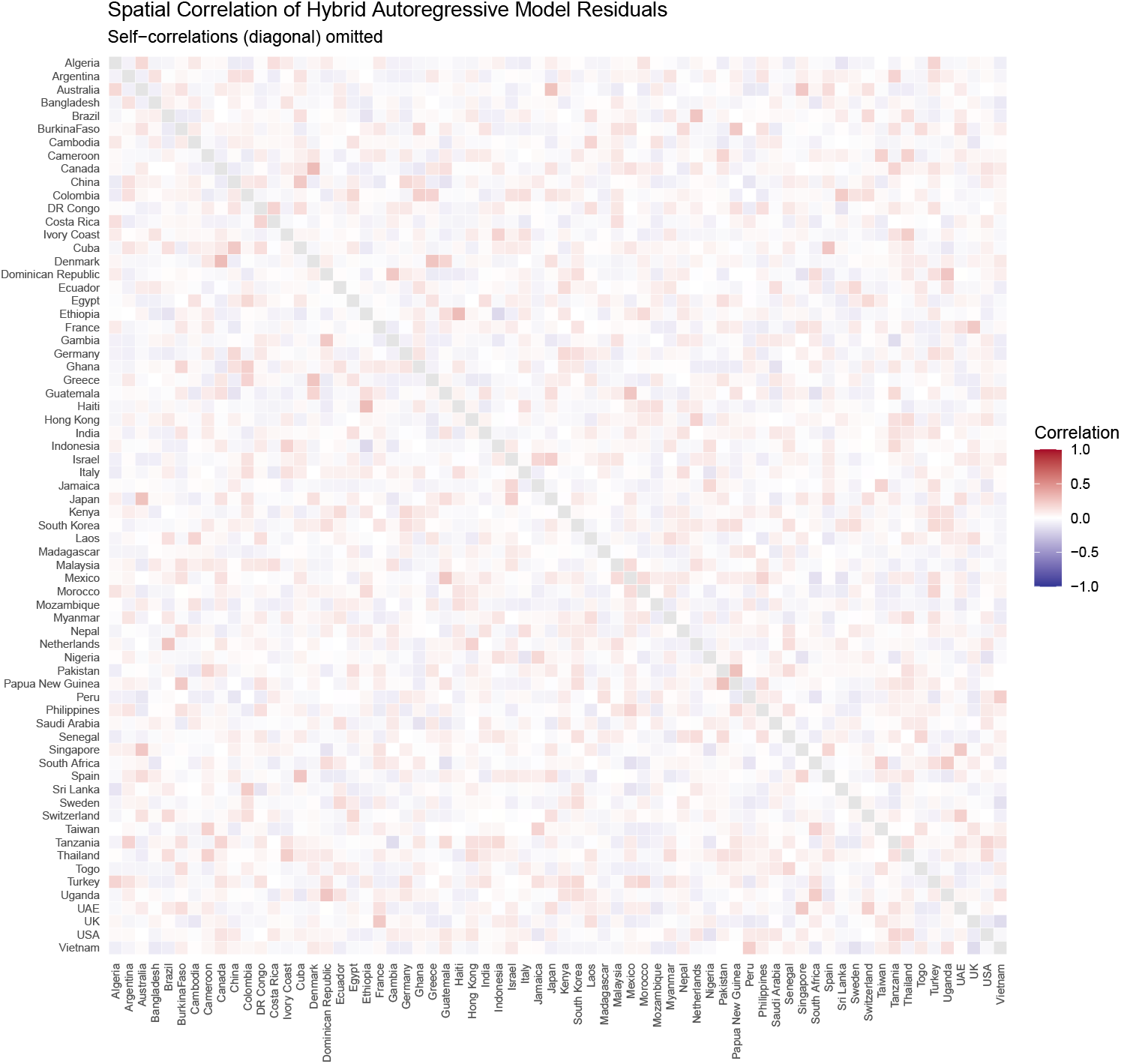
Residual spatial autocorrelation of the hybrid autoregressive model fitted to 2015-2019 data of all 68 countries. Conditions with differing expert classification as acute LRTIs are included.

### D.2 Sensitivity analysis: early identification of potential COVID-19 signals in 2020

**Fig. D11: Supplementary Figure S11.**
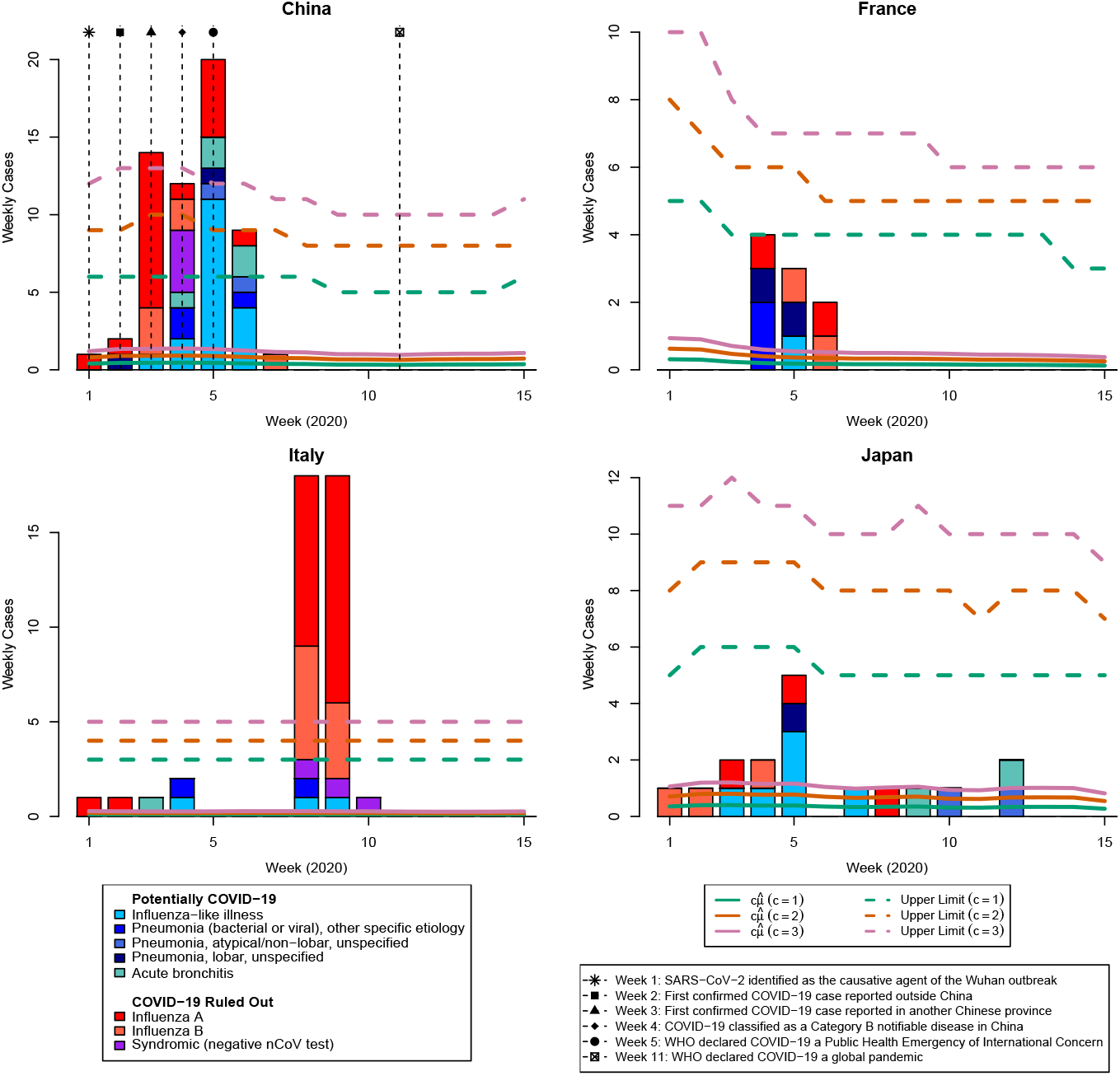
Number of weekly cases for travelers from China, France, Italy, and Japan, respectively, during the initial weeks of 2020, including conditions with differing expert classification as acute LRTIs. Robust Shewhart charts were set up based on the hybrid autoregressive model fitted to 2015–2019 data, and configured with various values for the maximum allowable multiplicative increase in travel volume under non-epidemic conditions (*c* = 1, 2, 3). The global significance level was set so that we expect approximately one false signal every 156 weeks (about three years). We applied a Bonferroni correction and, for 68 countries, set *α* = 9.4268 × 10^−5^, yielding an in-control average run length (ARL_0_) of approximately 10,000 weeks (about 192 years) at the individual country level. Solid lines show scaled mean predictions 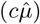, and dotted lines indicate the corresponding upper limits. Out of 68 countries, China and Italy were the only ones that triggered a signal under *c* = 1. France and Japan came close to signaling.

### D.3 Sensitivity analysis: goodness-of-fit (post-pandemic)

**Fig. D12: Supplementary Figure S12.**
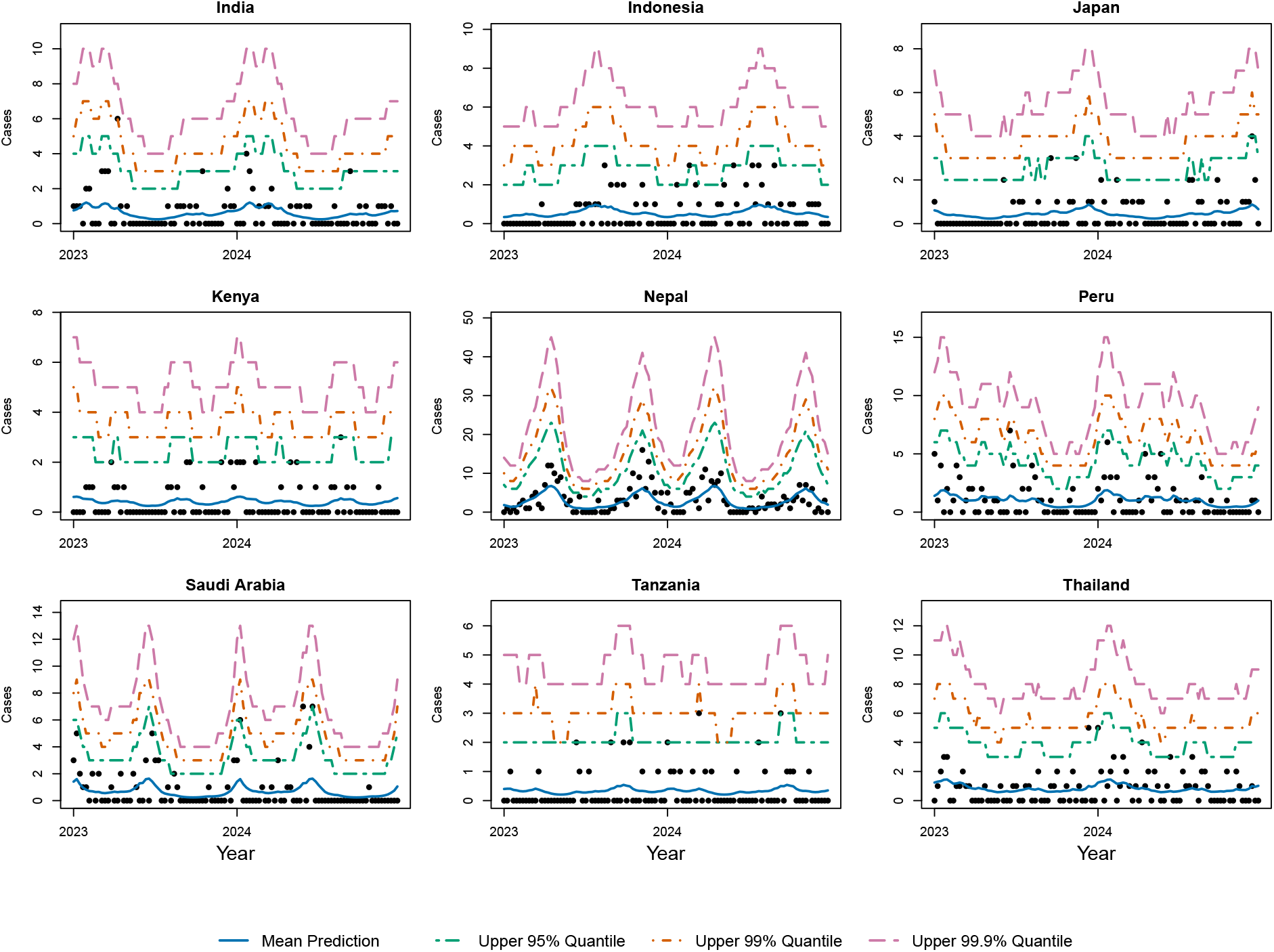
Number of weekly cases for the nine countries with the highest number of overall respiratory illnesses in the period 2023-2024 (India, Indonesia, Japan, Kenya, Nepal, Peru, Saudi Arabia, Tanzania, Thailand), including conditions with differing expert classification as acute LRTIs. The fit of the hybrid autoregressive model is visualized by the mean prediction 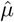 in solid blue, with upper 95%, 99% and 99.9% quantiles indicated by green, orange and purple dotted lines, respectively.

**Fig. D13: Supplementary Figure S13.**
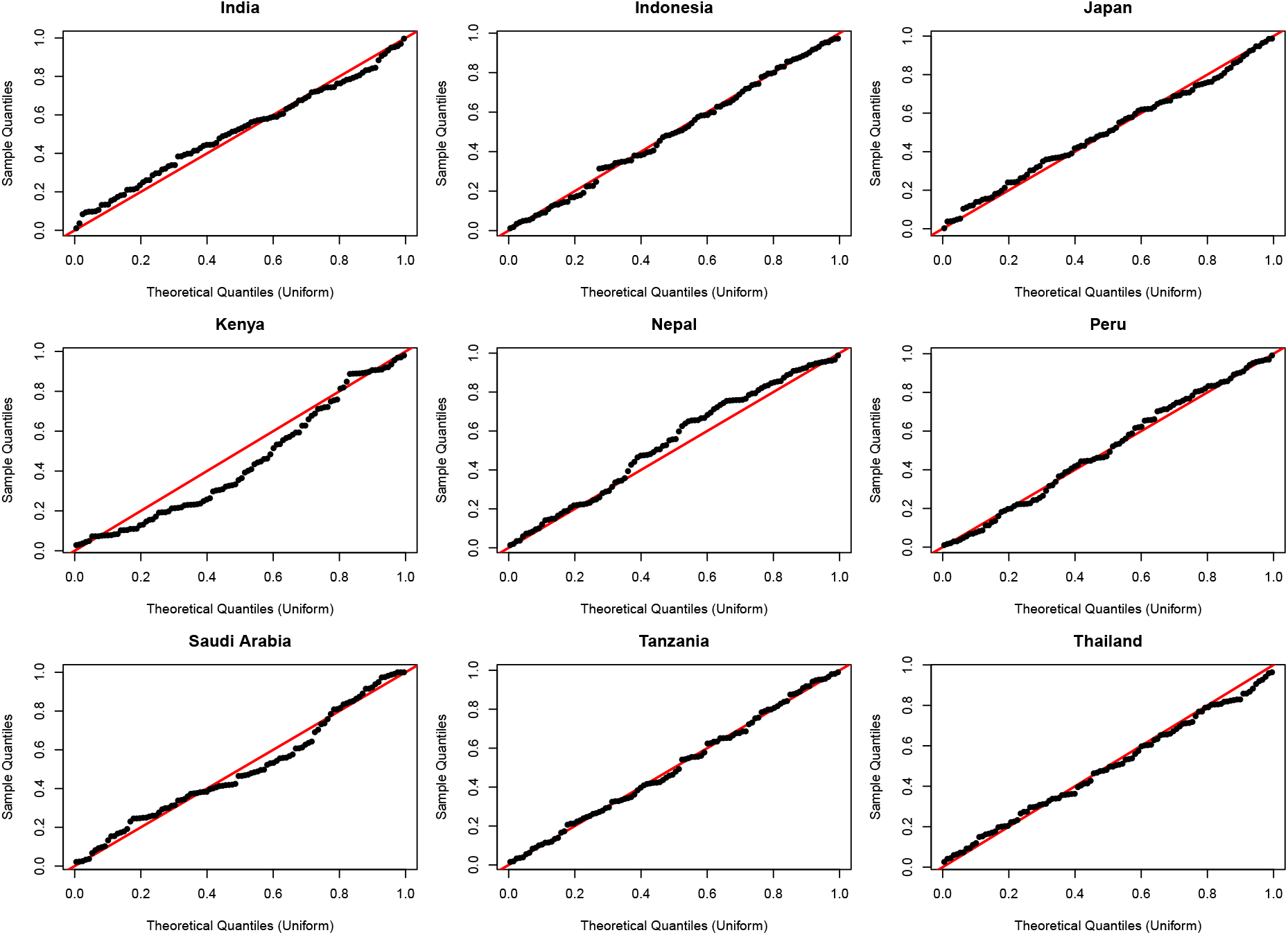
Quantile-Quantile residual plots of the hybrid autoregressive model fitted to data of nine countries with the highest number of overall respiratory illnesses in the period 2023-2024 (India, Indonesia, Japan, Kenya, Nepal, Peru, Saudi Arabia, Tanzania, Thailand), including conditions with differing expert classification as acute LRTIs.

**Fig. D14: Supplementary Figure S14.**
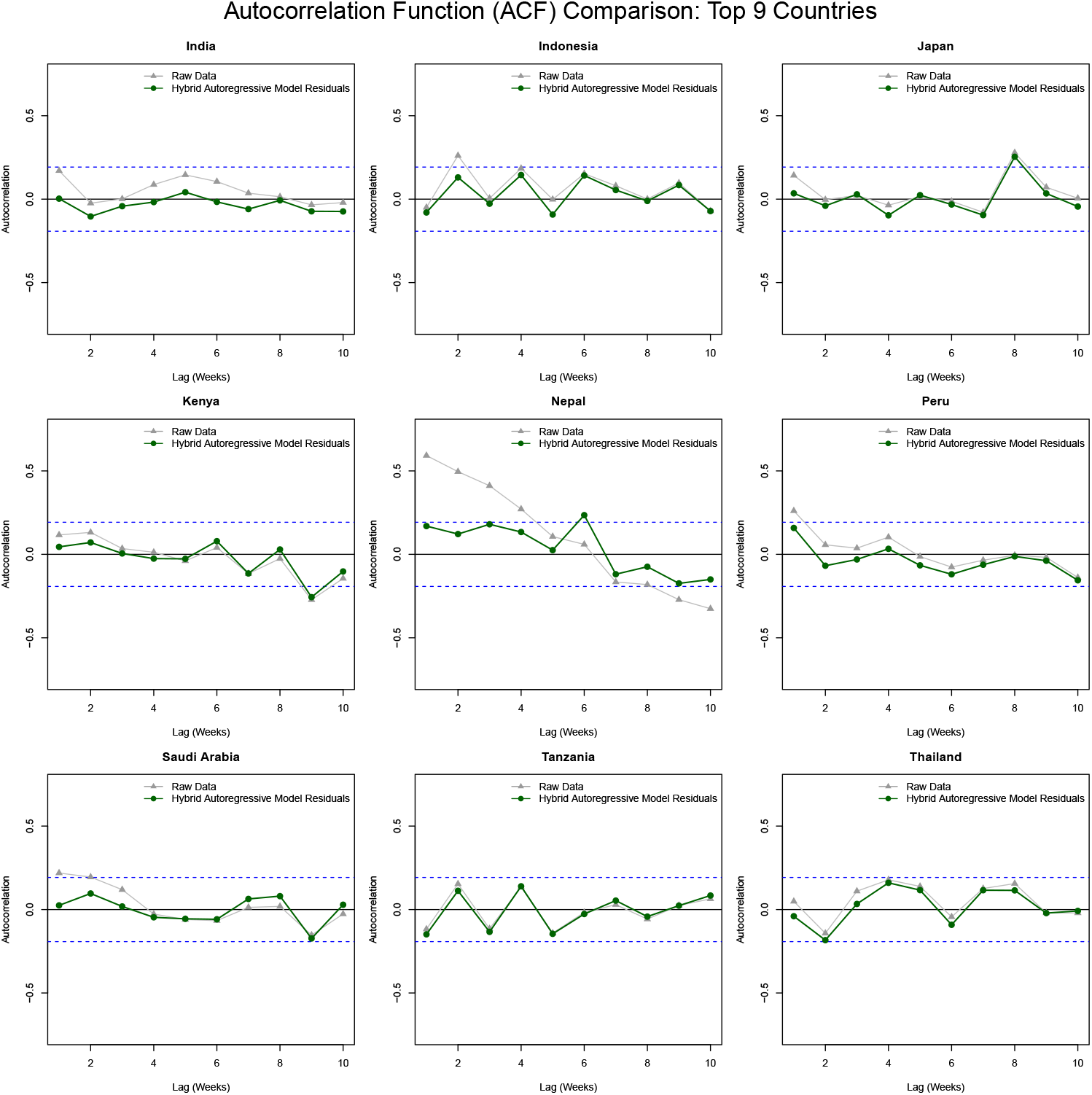
Residual temporal autocorrelation for lags 1-10 of the hybrid autoregressive model (green) fitted to data of the nine countries with the highest number of reported respiratory illnesses (including conditions with differing expert classification as acute LRTIs) in 2023-2024 (India, Indonesia, Japan, Kenya, Nepal, Peru, Saudi Arabia, Tanzania, Thailand), compared to the raw data ACF (grey). Blue dotted lines indicate 95% significance bounds 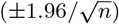.

**Fig. D15: Supplementary Figure S15.**
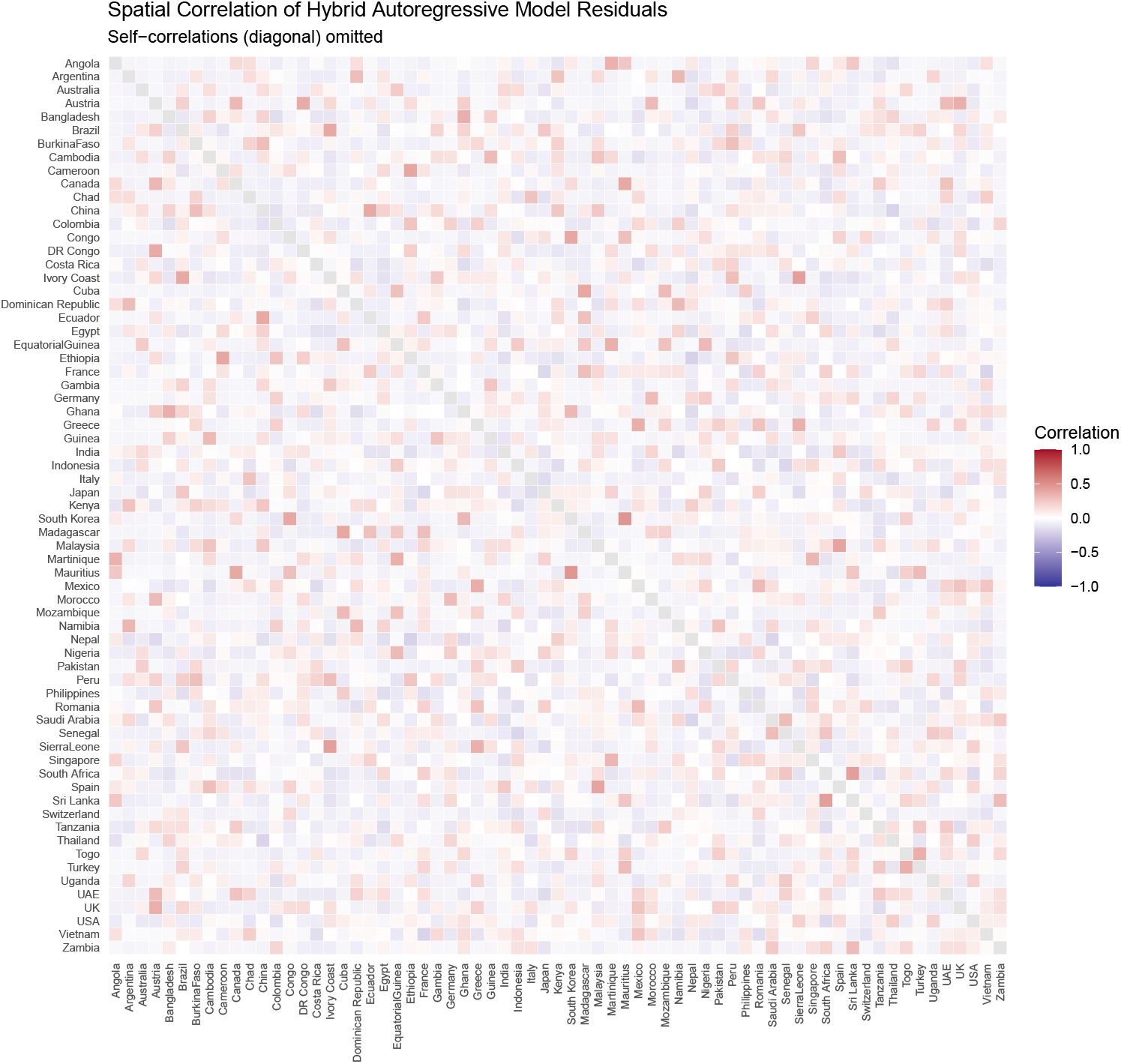
Residual spatial autocorrelation of the hybrid autoregressive model fitted to 2023-2024 data of all 67 countries. Conditions with differing expert classification as acute LRTIs are included.

## Footnotes

1 See 45 C.F.R. part 46; 21 C.F.R. part 56; 42 U.S.C. §241(d); 5 U.S.C. §552a; 44 U.S.C. §3501 et seq.

